# Single-value brain activity scores reflect both severity and risk across the Alzheimer’s continuum

**DOI:** 10.1101/2023.10.11.23296891

**Authors:** Joram Soch, Anni Richter, Jasmin M. Kizilirmak, Hartmut Schütze, Gabriel Ziegler, Slawek Altenstein, Frederic Brosseron, Peter Dechent, Klaus Fliessbach, Silka Dawn Freiesleben, Wenzel Glanz, Daria Gref, Michael T. Heneka, Stefan Hetzer, Enise I. Incesoy, Ingo Kilimann, Okka Kimmich, Luca Kleineidam, Elizabeth Kuhn, Christoph Laske, Andrea Lohse, Falk Lüsebrink, Matthias H. Munk, Oliver Peters, Lukas Preis, Josef Priller, Alfredo Ramirez, Sandra Roeske, Ayda Rostamzadeh, Nina Roy-Kluth, Klaus Scheffler, Matthias Schmid, Anja Schneider, Annika Spottke, Eike Jakob Spruth, Stefan Teipel, Jens Wiltfang, Frank Jessen, Michael Wagner, Emrah Düzel, Björn H. Schott

## Abstract

Single-value scores reflecting the deviation from (FADE score) or similarity with (SAME score) prototypical novelty-related and memory-related functional magnetic resonance imaging (fMRI) activation patterns in young adults have been proposed as imaging biomarkers of healthy neurocognitive aging. Here, we tested the utility of these scores as potential diagnostic and prognostic markers in Alzheimer’s disease (AD) and risk states like mild cognitive impairment (MCI) or subjective cognitive decline (SCD).

To this end, we analyzed subsequent memory fMRI data from individuals with SCD, MCI, and AD dementia as well as healthy controls (HC) and first-degree relatives of AD dementia patients (AD-rel) who participated in the multi-center DELCODE study (N = 468). Based on the individual participants’ whole-brain fMRI novelty and subsequent memory responses, we calculated the FADE and SAME scores and assessed their association with AD risk stage, neuropsychological test scores, CSF amyloid positivity, and ApoE genotype.

Memory-based FADE and SAME scores showed a considerably larger deviation from a reference sample of young adults in the MCI and AD dementia groups compared to HC, SCD and AD-rel. In addition, novelty-based scores significantly differed between the MCI and AD dementia groups. Across the entire sample, single-value scores correlated with neuropsychological test performance. The novelty-based SAME score further differed between Aβ-positive and Aβ-negative individuals in SCD and AD-rel, and between ApoE ε4 carriers and non-carriers in AD-rel.

Hence, FADE and SAME scores are associated with both cognitive performance and individual risk factors for AD. Their potential utility as diagnostic and prognostic biomarkers warrants further exploration, particularly in individuals with SCD and healthy relatives of AD dementia patients.

## Introduction

Cognitive decline and brain structural changes occur in most humans during aging, including in healthy individuals^1–3^. Explicit, and particularly, episodic memory, the ability to store, maintain, and retrieve single events^4^, is especially vulnerable to age-related decline, particularly in individuals at risk for Alzheimer’s disease (AD)^5–8^. However, inter-individual variability is high^9^, and distinguishing accelerated, but yet for-age normal cognitive decline from pre-clinical AD is challenging.

Mild cognitive impairment (MCI), defined as measurable cognitive decline with preserved functioning in activities of daily living^10,11^, is a well-characterized risk state for AD. Recently, subjective cognitive decline (SCD), defined by worry about deteriorating cognitive function despite normal performance, has been identified as a pre-MCI risk state^12,13^. Despite an increased risk of developing AD dementia compared to the general population, not all individuals with MCI and even fewer with SCD progress to dementia. Therefore, the establishment of biomarkers reflecting an individual’s risk for AD dementia is highly desirable^14–17^.

Currently, *loco typico* brain structural changes in AD have yielded several neuroimaging biomarkers for AD, including reduced gray matter volume (GMV)^18,19^, reduced hippocampal volumes^20^, and white matter lesion load^21,17^. Moreover, memory-related functional magnetic resonance imaging (fMRI) may constitute a helpful measure for differentiating normal from at-risk neurocognitive aging^22–25^.

In the commonly employed subsequent memory paradigm, participants encode stimuli, which they are subsequently asked to recall or recognize. Successful encoding, assessed via comparison of subsequently remembered vs. forgotten items (i.e., subsequent memory effect), typically elicits increased activations of the bilateral medial temporal lobe (MTL), including the hippocampus, as well as inferior temporal, parieto-occipital and prefrontal cortices (for meta-analyses, see ^25,26^). Presenting pre-familiarized stimuli intermixed with novel stimuli during encoding additionally allows the study of novelty effects (i.e., novel vs. familiar items^27,22^), which typically encompass activations in MTL regions and deactivations of default mode network (DMN) regions like the precuneus^28–30^.

Despite the relatively large number of studies on memory encoding in AD and MCI (for meta-analyses, see ^31–34^), only few studies have reported actual subsequent memory effects^35–37^. Instead, most studies report on encoding compared to a low-level baseline or on novelty effects^31,34,24^. One reason for this may be that poor episodic memory in AD, and to some extent in MCI, reduces the signal-to-noise ratio of encoding-specific fMRI responses, making it difficult to differentiate between subsequently remembered and forgotten items. Compatibly, we have recently shown that, when comparing first-level fMRI models using Bayesian model selection, memory-invariant fMRI models provide a better fit than subsequent memory models in individuals with MCI or mild AD dementia^38^.

In previous studies investigating healthy older adults^22,29,30,39^, single-value scores extracted from whole-brain fMRI contrast maps for novelty processing and subsequent memory have been proposed as potential biomarkers of neurocognitive aging. Single-value scores quantify *F*unctional *A*ctivity *D*eviation during *E*ncoding (FADE) or *S*imilarity of *A*ctivations during *M*emory *E*ncoding (SAME) in relation to prototypical activations in young adults. Thus, these scores provide reductionist measures of an individual’s memory network integrity. In a sample of healthy young and older adults, we have previously reported that these scores differed between age groups, correlated with memory performance^39,30^, and were robust against potential confounds like MRI scanner or reference sample^29^.

Here, we investigated to what extent FADE and SAME reflect neurocognitive decline across the AD risk spectrum. In addition to psychometric tests of memory performance and functional neuroimaging, we examined the effects of the well-established ApoE genetic risk factor and of the Aβ 42/40 ratio in cerebrospinal fluid (CSF)^40–43^. We applied our previously described approach^29^ to a large cohort from the DZNE Longitudinal Cognitive Impairment and Dementia Study (DELCODE)^44^, including healthy controls (HC), individuals with SCD, MCI, and mild AD dementia, and first-degree relatives of AD dementia patients (AD-rel).

We hypothesized that FADE and SAME scores would be affected by clinical severity across the AD risk spectrum, with increasing FADE scores (i.e., larger deviation from prototypical activation patterns in a reference sample of young adults) and decreasing SAME scores (i.e., lower similarity with activation patterns in the reference sample). We further hypothesized that (i) acquisition site, gender and educational status would not significantly affect the scores^29^; (ii) the scores would correlate with episodic memory performance and additional cognitive measures across participant groups^30^; and that (iii) ApoE ε4 allele carriage and amyloid positivity (as determined by the CSF Aβ42/40 ratio), would be associated with higher FADE and lower SAME scores within or across diagnostic groups.

## Materials and methods

### Study cohort

The study sample consisted of participants from the DELCODE Study (https://www.dzne.de/en/research/studies/clinical-studies/delcode/)^44^, including individuals with SCD, MCI or early-stage AD as well as cognitively unimpaired older control participants and healthy first-degree relatives of patients with AD dementia. DELCODE is a multi-center memory clinic-based study focusing on preclinical stages of AD, conducted across different sites of the German Center for Neurodegenerative Diseases (DZNE).

Complete baseline data (i.e., data from the first study visit) was available for 844 participants. We excluded participants (i) without available diagnosis, (ii) missing or incomplete fMRI data, and (iii) missing essential meta-data, resulting in a final sample size of N = 468 (HC: 128; SCD: 199; MCI: 74; AD: 21; AD-rel: 46). Participant demographics are reported in Table 1.

**Table 1.**
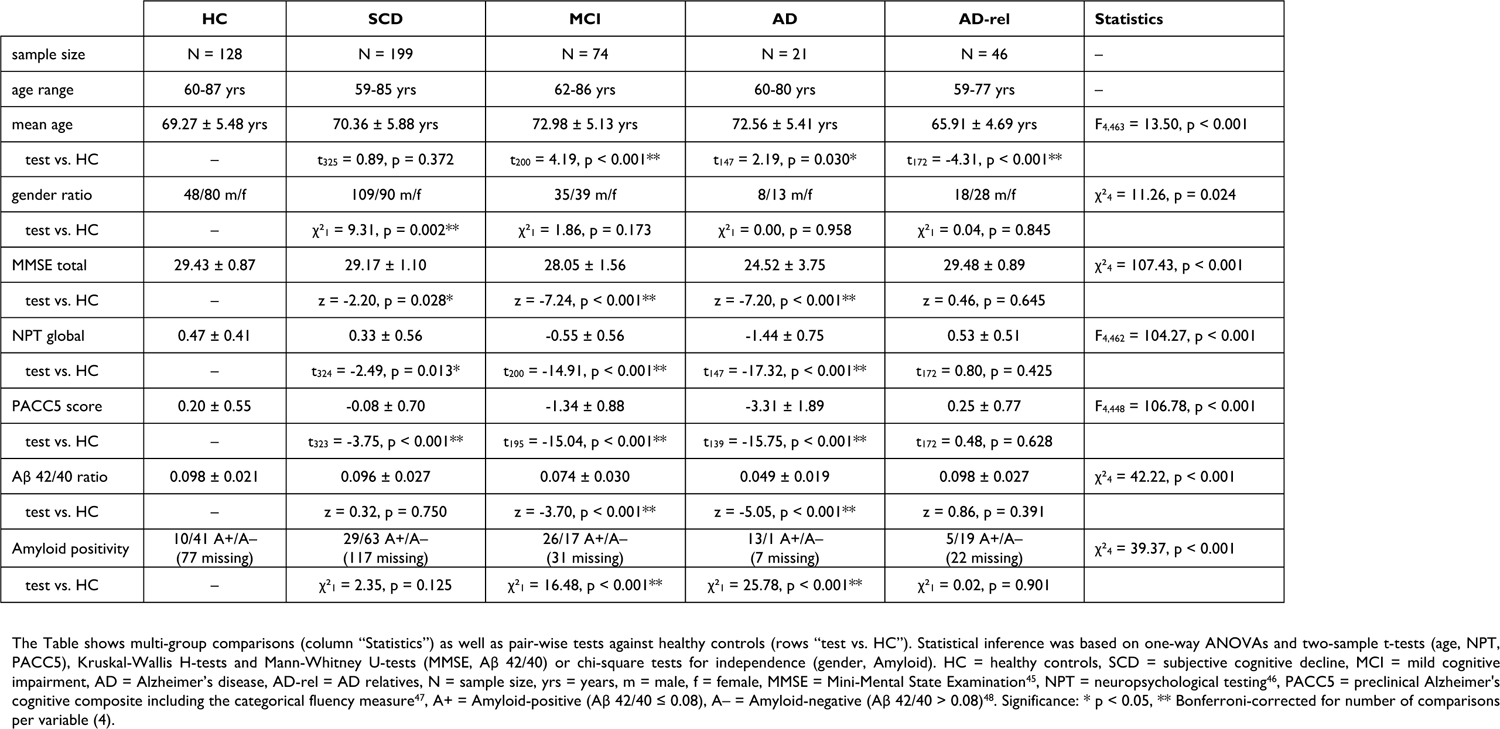
Demographic information of participant groups.

### Methods overview

Apart from using a different study cohort, comprising five (HC, SCD, MCI, AD and AD-rel) rather than two (healthy young and older adults) groups and the multi-centric acquisition, the present study employed the same MRI acquisition parameters, fMRI processing pipeline and analysis protocols as in ^29^. The neuropsychological test batteries differed, owing to the demographics and clinical characteristics of the study samples. All data analyses were performed after publication of the reference study^29^ (Table S2), following the approval of the analysis protocol by the DELCODE steering committee. The corresponding data analysis proposal is available from the authors upon request.

### Experimental paradigm

Participants performed an adapted version of a previously described memory encoding task^22^ as part of the DELCODE study protocol^49,50^, which was also employed in our earlier study^29^. Briefly, participants viewed photographs of indoor and outdoor scenes, which were either novel at the time of presentation (i.e., 44 indoor and 44 outdoor scenes) or repetitions of two pre-familiarized “master” images (i.e., 22 indoor and 22 outdoor trials). In a recognition memory test 70 minutes later, participants were shown all novel images from the encoding session, now considered “old” stimuli (88 images in total), and previously unseen, that is, “new” stimuli (44 images in total). Participants were asked to provide a recognition-confidence rating for each image, using a five-point Likert scale ranging from “sure new” (1) over “don’t know” (3) to “sure old” (5).

### MRI data acquisition

MRI data were acquired at eight different sites across Germany using Siemens 3T MR tomographs. All sites followed the DELCODE MRI protocol^29,44,49^. Structural MRI included a T1-weighted MPRAGE image (voxel size = 1 x 1 x 1 mm) and phase and magnitude fieldmaps for later spatial artifact correction. Functional MRI consisted of 206 T2*-weighted echo-planar images (EPIs; TR = 2.58 s, voxel size = 3.5 x 3.5 x 3.5 mm) acquired during the encoding session of the memory task (09:01 min), and a resting-state session (180 scans, not used here).

### MRI data processing

Data processing and analysis were performed using Statistical Parametric Mapping, version 12 (SPM12; Wellcome Centre for Human Neuroimaging, University College London, London, UK; https://www.fil.ion.ucl.ac.uk/spm/software/spm12/) and in-house MATLAB scripts (https://github.com/JoramSoch/FADE_SAME). Preprocessing of fMRI data included correction for acquisition time (*slice timing*), head motion (*realignment*), and magnetic field inhomogeneities using the fieldmaps (*unwarping*), coregistration of the T1-weighted MPRAGE image to the mean EPI computed during realignment, segmentation of the coregistered MPRAGE image, subsequent normalization of unwarped EPIs into the MNI standard space (voxel size = 3 x 3 x 3 mm), and spatial smoothing of the normalized EPIs (FWHM = 6 mm).

Statistical analysis of the fMRI data was based on voxel-wise general linear models (GLMs) that included two onset regressors, representing novel images (*novelty* regressor) and master images (*master* regressor), six head motion regressors obtained from realignment and a constant representing the implicit baseline. The novelty regressor was parametrically modulated with the arcsine-transformed subsequent memory response, yielding a regressor reflecting encoding success (Appendix, eq. 1). This model (“GLM_1t-a”, cf. Table 3 in ^38^) had emerged as the winning theoretical parametric GLM from Bayesian model selection between fMRI models in an independent cohort of healthy young and older adults^28^, as well as in the HC, SCD and AD-rel groups from the DELCODE study^38^.

### Single-value fMRI scores

fMRI contrast maps for novelty processing (novel vs. master images) and subsequent memory (parametric memory regressor) were calculated for each subject (Supplementary Methods). From both contrasts, two single-value fMRI scores were computed: FADE^22^ and SAME^29^. The FADE score is calculated as the average t-value of an older participant on a specific contrast in all voxels in which young participants show a positive effect on this contrast, subtracted from the average t-value of the same contrast outside those voxels (Appendix, eq. 2). The SAME score is calculated as the average of reduced activations of an older individual in all voxels in which young adults show a positive effect, plus the average of reduced deactivations in all voxels with a negative effect (Appendix, eq. 3).

In addition to the directionality, the SAME scores differ from the FADE scores by:

1. their (semi-)quantitative nature as they reflect voxel-wise differences between the subject’s and reference sample’s parameter estimates rather than the average t-values inside vs. outside a binarized activation mask,
2. explicitly considering deactivations, particularly in default mode network (DMN) regions (cf. Fig. 1a in ^29^), which may reflect early disturbances of memory network integrity in individuals, particularly in individuals with SCD^24^, and
3. accounting for the variance within the reference sample (Appendix, eq. 3).

**Figure 1.**
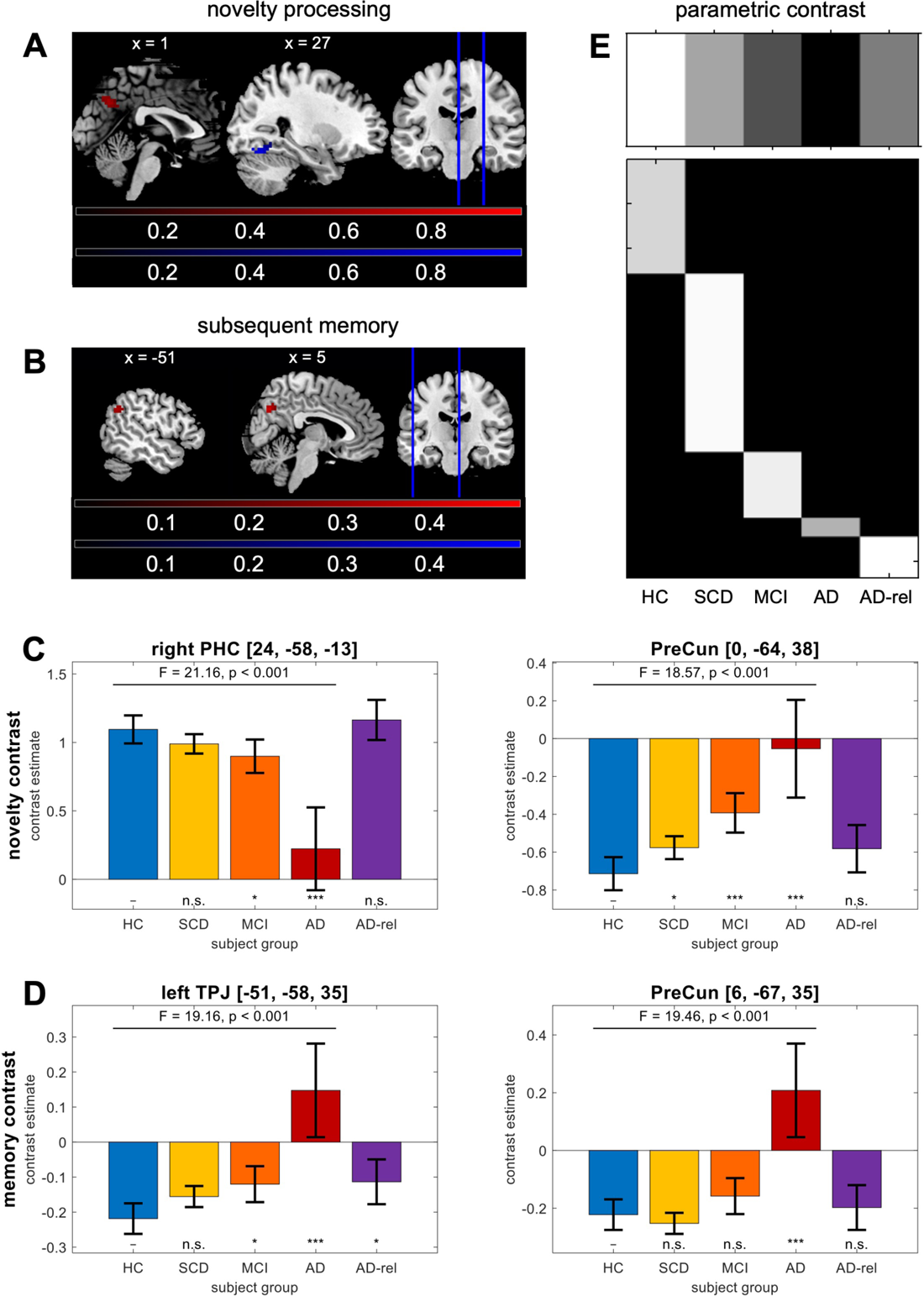
Diagnosis-related activation differences in the human memory network. Encoding-related fMRI activity was compared across five diagnosis groups (HC, SCD, MCI, AD, AD-rel). Brain sections show significant effects of disease severity for **(A)** the novelty contrast (novel vs. master images) and **(B)** the memory contrast (subsequent memory regressor), obtained using **(E)** a parametric F-contrast (c = [+3, +1, –1, –3, 0]) testing for a linear decrease or increase with disease progression (excluding AD relatives, because they cannot be meaningfully included into the rank order of AD risk stages). Voxel colors indicate average differences between healthy controls and Alzheimer’s patients, resulting from either higher activity in disease (AD > HC, red) or higher activity in health (HC > AD, blue). Bar plots show group-level contrast estimates and 90% confidence intervals for **(C)** the novelty contrast (novel vs. master images) and **(D)** the memory contrast (subsequent memory regressor), extracted from the local maxima in A and B. Statistics inside the panels correspond to **(E)** an F-contrast testing for a parametric increase or decrease with disease severity (F/p-values; all F-values are F_1,463_ statistics) and t-contrasts testing each group against healthy controls (significance markers). Abbreviations: HC = healthy controls, SCD = subjective cognitive decline, MCI = mild cognitive impairment, AD = Alzheimer’s disease, AD-rel = AD relatives. Significance: * p < 0.05, Bonferroni-corrected for ** number of tests per region (4) or *** number of tests and number of regions (4 x 2).

In a previous study on the neuropsychological correlates of the single-value scores, we have shown that, despite FADE and SAME being negatively correlated, there are relationships with cognitive performance measures unique to either FADE or SAME scores^30^. For more information on the calculation and interpretation of the scores, see the original descriptions (cf. Fig. 1 and Appendix A in ^29^).

### Psychometric testing

Memory performance in the fMRI task was measured as “A-prime”, the area under the curve in a receiver-operating characteristic (ROC) analysis of the subsequent memory reports (cf. Appendix B in ^29^).

Participants completed a battery of neuropsychological tests. The Mini-Mental State Examination (MMSE) score^45,52,53^ was a main criterion for the diagnosis of MCI and mild AD. The preclinical Alzheimer cognitive composite score (PACC5) is derived as a composite measure based on the following neuropsychological test scores:

1. the Total Recall score from the Free and Cued Selective Reminding Test (FCSRT)^54^,
2. the Delayed Recall score on the Logical Memory IIa subtest from the Wechsler Memory Scale (WMS)^55^,
3. the Digit Symbol Substitution Test score from the Wechsler Adult Intelligence Scale– Revised (WAIS-R)^56^,
4. the MMSE total score, and
5. category fluency as a measure of semantic knowledge^47^.

For each subject, the PACC5 was defined as the sum of all z-transformed values from each sub-score^57^.

The neuropsychological test (NPT) score represents the mean score of five factors derived from a factor analysis conducted on a large variety of neuropsychological tests^46^. These include components of the PACC5 and several subscales from the FCSRT, the Trail-Making Test, Clock Drawing Test, additional WMS subscales (Logical Memory 1 and 2), the Face Naming Test, Symbol digit modalities test, Boston Naming Task, and Flanker Task.

### Fluid biomarkers

Amyloid-beta and tau epitopes in CSF (Aβ-42/40 ratio, total Tau, p-Tau181) were determined using commercially available kits according to vendor specifications: V-PLEX Aβ Peptide Panel 1 (6E10) Kit (K15200E) and V-PLEX Human Total Tau Kit (K151LAE) (Mesoscale Diagnostics LLC, Rockville, USA), and Innotest Phospho-Tau (181P) (81581) (Fujirebio Germany GmbH, Hannover, Germany). For more details on CSF biomarkers, see previous DELCODE publications (e.g., ^23^).

Genotypes of rs7412 and rs429358, the single nucleotide polymorphisms defining the ApoE ε2, ε3, and ε4 alleles, were identified using commercially available TaqMan® SNP Genotyping Assay (ThermoFisher Scientific; for details, see previous DELCODE publications, e.g. ^44^).

### Statistical analyses

The goal of the present analyses was two-fold: First, we aimed to assess the robustness of FADE and SAME scores against confounding variables (e.g., Table 2). Second, we aimed to assess potential relationships of the scores with factors previously implicated in cognitive aging or increased risk for developing AD dementia (e.g., Figure 5).

**Table 2.**
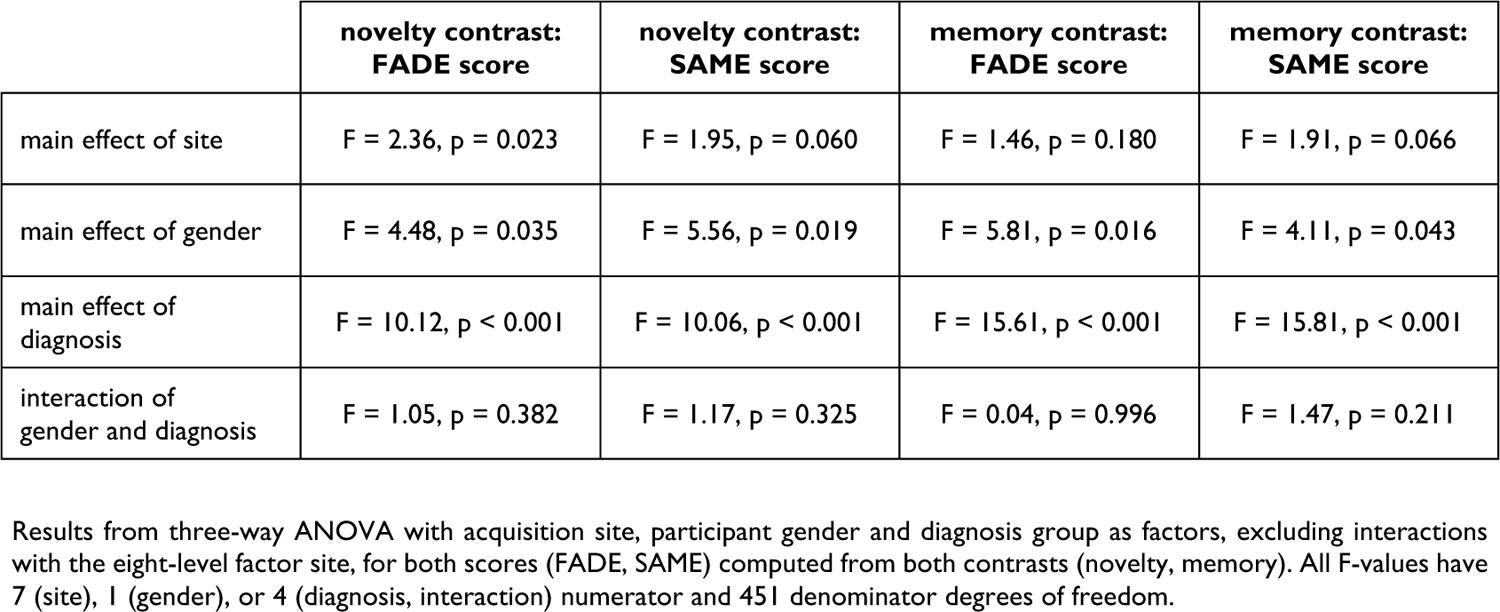
Effects of site, gender and diagnosis on fMRI scores.

|  | novelty contrast:<br>FADE score | novelty contrast:<br>SAME score | memory contrast:<br>FADE score | memory contrast:<br>SAME score |
| --- | --- | --- | --- | --- |
| main effect of site | F = 2.36, p = 0.023 | F = 1.95, p = 0.060 | F = 1.46, p = 0.180 | F = 1.91, p = 0.066 |
| main effect of gender | F = 4.48, p = 0.035 | F = 5.56, p = 0.019 | F = 5.81, p = 0.016 | F = 4.11, p = 0.043 |
| main effect of diagnosis | F = 10.12, p < 0.001 | F = 10.06, p < 0.001 | F = 15.61, p < 0.001 | F = 15.81, p < 0.001 |
| interaction of gender and diagnosis | F = 1.05, p = 0.382 | F = 1.17, p = 0.325 | F = 0.04, p = 0.996 | F = 1.47, p = 0.211 |

Results from three-way ANOVA with acquisition site, participant gender and diagnosis group as factors, excluding interactions with the eight-level factor site, for both scores (FADE, SAME) computed from both contrasts (novelty, memory). All F-values have 7 (site), 1 (gender), or 4 (diagnosis, interaction) numerator and 451 denominator degrees of freedom.

**Table 3.** Comparison of ApoE genotypes to population distribution.

| Group | $\epsilon 2/\epsilon 2$ | $\epsilon 2/\epsilon 3$ | $\epsilon 2/\epsilon 4$ | $\epsilon 3/\epsilon 3$ | $\epsilon 3/\epsilon 4$ | $\epsilon 4/\epsilon 4$ | Statistics |
| --- | --- | --- | --- | --- | --- | --- | --- |
| Population | 0.60 | 12.46 | 2.81 | 59.80 | 22.21 | 2.11 | – |
| HC (N = 125) | 0.80 | 15.20 | 0.80 | 64.00 | 16.80 | 2.40 | $\chi^2_2 = 1.78$, p = 0.410 |
| SCD (N = 193) | 1.04 | 11.92 | 2.59 | 54.40 | 27.98 | 2.07 | $\chi^2_3 = 3.58$, p = 0.310 |
| MCI (N = 73) | 2.74 | 5.48 | 5.48 | 45.21 | 31.51 | 9.59 | $\chi^2_2 = 11.26$, p = 0.004 |
| AD (N = 21) | 0.00 | 4.76 | 0.00 | 14.29 | 61.90 | 19.05 | $\chi^2_1 = 36.59$, p < 0.001 |
| AD-rel (N = 46) | 0.00 | 6.52 | 2.17 | 63.04 | 23.91 | 4.35 | $\chi^2_2 = 1.87$, p = 0.393 |

To investigate the robustness and stability of the scores, FADE and SAME scores were (i) subjected to between-subject ANOVAs using site, gender and diagnostic group as factors, (ii) analyzed with score-wise mixed ANOVAs using diagnosis and contrast as factors, and (iii) computed based on different reference samples.

To investigate relationships between the scores and variables relevant for cognitive aging, FADE and SAME scores were analyzed as a function of (i) baseline diagnosis, (ii) chronological age, (iii) memory performance in the fMRI task, (iv) educational and employment years, (v) demographic/lifestyle factors like BMI, (vi) neuropsychological test scores such as MMSE, NPT and PACC5, (vii) fluid biomarkers (total-tau, p-Tau181, Aβ-42/40 ratio), and the categorical variables (viii) amyloid positivity, (ix) ApoE genotype, and (x) educational status.

In total, these investigations resulted in ten statistical analyses (Table S1). All analyses, except for the mixed ANOVAs, were conducted and are reported separately for each combination of contrast and score (i.e., for all four types of scores: novelty-FADE, novelty-SAME, memory-FADE, memory-SAME; e.g., Figure 2).

**Figure 2.**
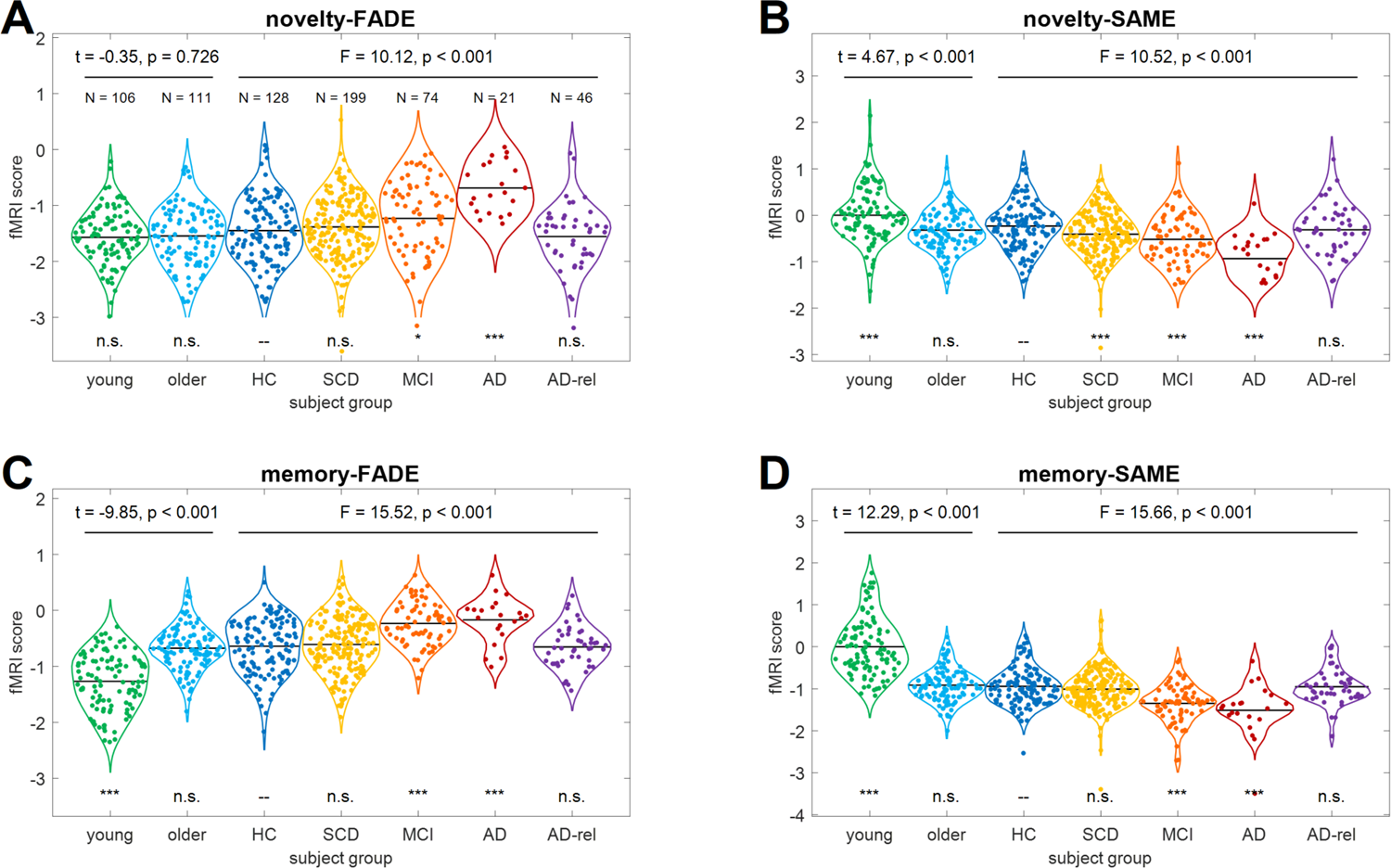
FADE and SAME scores as a function of fMRI contrast, score type and diagnostic group. Single-value fMRI scores are shown as violin and sina plots for **(A)** the FADE score and **(B)** the SAME score computed from the novelty contrast as well as **(C)** the FADE score and **(D)** the SAME score computed from the memory contrast. Scores were calculated for young (green) and older (light blue) subjects from the original study as well as HCs (dark blue), individuals with SCD (yellow), MCI patients (orange), AD patients (red) and AD relatives (violet) from the DELCODE study. Sample sizes are given in the upper-left panel. Horizontal bars correspond to group-wise means. Statistics inside the panels correspond to a two-sample t-test between young and older adults (t/p-value; 215 degrees of freedom, DOF), a one-way ANOVA across DELCODE diagnostic groups (F/p-value; 4 numerator and 463 denominator DOFs) and two-sample t-tests of each group against DELCODE healthy controls (significance markers). Abbreviations: FADE = functional activity deviation during encoding, SAME = similarities of activations during memory encoding, HC = healthy controls, SCD = subjective cognitive decline, MCI = mild cognitive impairment, AD = Alzheimer’s disease, AD-rel = AD relatives. Significance: * p < 0.05, Bonferroni-corrected for ** number of tests per score (6) or *** number of tests and number of scores (6 x 4). This figure extends results reported earlier (see Fig. 3 in ^29^).

### Predictive analyses

To assess the predictive utility of the single-value fMRI scores, we performed support vector machine (SVM) classification analyses, using all four scores as features and grouping the entire sample into several distinct subgroups, based on, for example, diagnostic group, ApoE genotype or Amyloid status (Table S4).

In each classification analysis, SVMs were calibrated with regularization hyperparameter C = 1 and using k = 10-fold cross-validation. To account for unequal sample sizes among participant groups, we repeatedly drew subsamples with a constant number of observations per class. Classification accuracy and 90% confidence interval as measures of predictive performance were obtained as averages across all S = 1000 subsamples. All predictive analyses were implemented using Machine Learning for MATLAB (https://github.com/JoramSoch/ML4ML).

## Results

### Novelty- and memory-related fMRI responses across diagnostic groups

We first investigated the voxel-wise differences between participant groups with respect to novelty and memory contrasts. To this end, we computed a second-level one-way ANOVA in SPM with diagnostic group (HC, SCD, MCI, AD, AD-rel) as between-subjects factor, and thresholded the statistical map for a parametric effect of diagnosis (Figure 1E), corrected for family-wise error (FWE) at cluster level (cluster-defining threshold, CDT = 0.001, extent threshold k = 35 (novelty) and k = 32 (memory); cf. ^58^).

We found significant effects on both fMRI contrasts (Figure 1A/B), implicating brain regions previously implicated in (visual) episodic memory formation (Figure S1), including MTL regions like parahippocampal cortex (PHC) and hippocampus as well as the precuneus (PreCun) and the temporo-parietal junction (TPJ).

Closer inspection of the activation patterns across participant groups (Figure 1C/D) revealed that (i) some of these differences were based on reduced activations in AD risk states compared to HC, especially in regions belonging to the human memory network (e.g. novelty: right PHC, Figure 1C); and (ii) some of these effects resulted from reduced deactivations in AD disease states compared to HC, especially in DMN regions (e.g. memory: left TPJ and PreCun, Figure 1D).

### FADE and SAME scores across the AD risk spectrum

When comparing the single-value fMRI scores across participant groups, we observed three tendencies. First, differences between healthy young and older participants replicate earlier results^29^, with significant effects of age group for all scores except for the FADE score from the novelty contrast (Figure 2A). As the DELCODE study did not include young participants, comparisons with young adults were conducted with the young participants from ^29^ (for details, see Supplementary Methods). Second, nominal differences largely mirrored the stages of the AD risk spectrum, with increasing risk being associated with more atypical fMRI scores (SAME scores: young > older ≈ HC ≈ AD-rel > SCD > MCI > AD; FADE scores: reverse order; Figure 2). Third, there were no significant differences between older subjects from ^29^, healthy controls from the DELCODE study, and AD relatives from the DELCODE study for any of the scores. SCD and healthy participant groups (HC, AD-rel) only differed in the novelty-SAME score (Figure 2B).

Memory-based scores did not significantly differ between the MCI and AD groups (FADE: t_93_ = −0.67, p = 0.504; SAME: t_93_ = 1.34, p = 0.182). They did, however, significantly differentiate both groups from all other diagnostic groups (FADE: t_466_ = −5.57, p < 0.001.; SAME: t_466_ = 5.46, p < 0.001; two-sample t-test for HC/SCD/AD-rel vs. MCI/AD; Figure 2C/D). Novelty-based scores did not significantly differ between the MCI and SCD groups (FADE: t_271_ = −1.90, p = 0.058; SAME: t_271_ = 1.66, p = 0.099). They did, however significantly differentiate the MCI and AD groups (FADE: t_93_ = −3.52, p < 0.001; SAME: t_93_ = 3.05, p = 0.003; Figure 2A/B).

When comparing novelty and memory contrasts, holding score type constant, we found significant interactions of diagnosis and contrast for both scores (FADE: F_4,463_ = 18.78, p < 0.001; SAME: F_4,463_ = 19.80, p < 0.001; Supplementary Results and Table S3).

### Robustness and stability of FADE and SAME scores

To control for potential confounding variables, we computed a three-way between-subjects ANOVA to assess effects of (i) acquisition site (8 sites; cf. Table 1 in ^38^), (ii) gender (male vs. female), and (iii) diagnostic group (HC, SCD, MCI, AD, AD-rel). Because the factor site had eight levels, we did not include interactions with site in this model. For detailed statistics, see Table 2.

The main effect of site was significant for the novelty-FADE score, but not when correcting for multiple comparisons (uncorrected p = 0.023). The main effect of gender was significant for all four scores, reflecting higher FADE scores and lower SAME scores in men compared to women (Figure S2), but not when correcting for multiple comparisons (uncorrected p-values in range 0.016 < p < 0.043). Main effects of diagnostic group remained significant for all scores when controlling for site and gender. There were no interactions between gender and diagnostic group.

Importantly, in addition to their robustness to gender and acquisition site, the scores were also stable when using a different, independent sample of young adults as reference (Supplementary Methods, Results, Table S5, Figures S3 and S4).

### FADE and SAME scores correlate with indices of cognitive aging and AD risk

To identify associations of the scores with indices of cognitive aging beyond diagnostic group, we computed partial correlations between the scores (novelty/memory x FADE/SAME) and markers of cognitive functioning (e.g., memory performance) lifestyle or demographic factors (e.g., BMI, education), and neurochemical and genetic markers (e.g., Aβ-42/40 ratio). To account for diagnostic group (HC, SCD, MCI, AD, AD-rel), we computed the correlations between residual independent variables and residual fMRI scores after removing group-wise means from both, correcting for multiple comparisons.

These partial correlations revealed several patterns (Figure 3): FADE and SAME scores (i) show significant correlations with chronological age, mainly supported by the large SCD group (Supplementary Methods, Results, Figures S7 and S8), (ii) correlate significantly with memory performance in the fMRI task (A-prime; Section “Psychometric testing”), (iii) are not significantly correlated with lifestyle-driven factors such as educational (for details, see Supplementary Methods, Results, and Figure S6) and employment years as well as height, weight and BMI; (iv) show weakly significant correlations with MMSE and stronger significant correlations with NPT and PACC5 scores, and, finally, (v) there is weak evidence for an association with total tau and phospho-tau and robust evidence for an association with the Aβ 42/40 ratio, but only for novelty-based scores.

**Figure 3.**
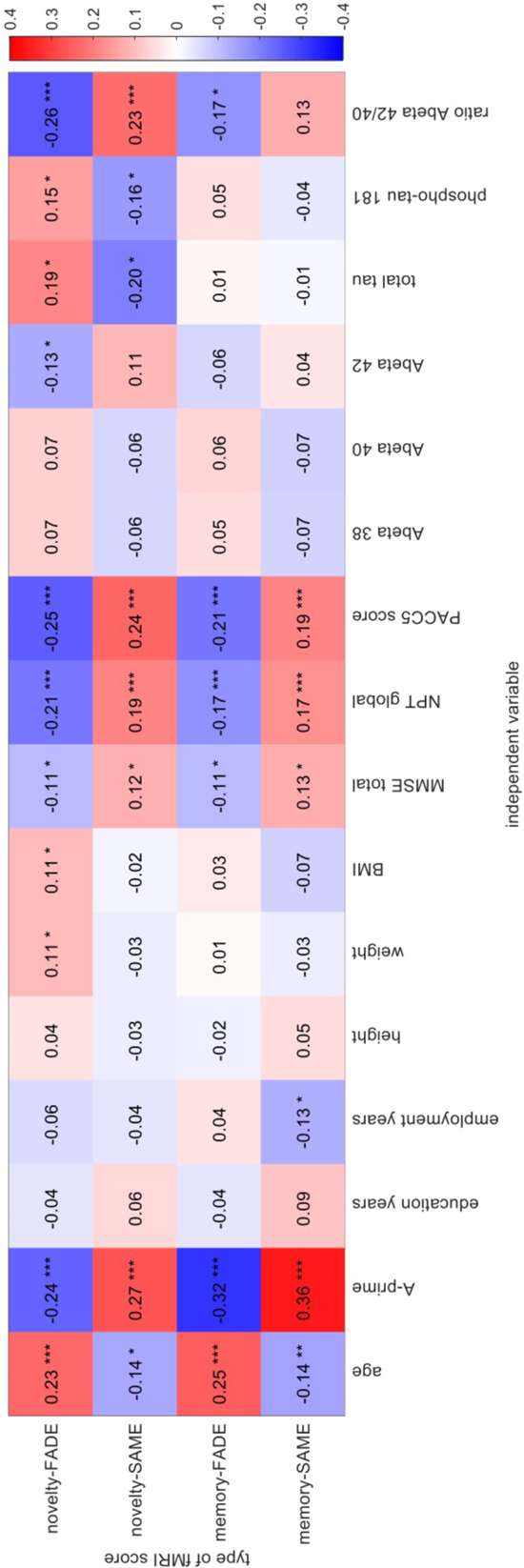
Partial correlations of FADE and SAME scores with other indices of cognitive aging. Positive (red) and negative (blue) partial correlations of single-value fMRI scores (y-axis) with selected independent variables (x-axis), accounting for participant group membership. Abbreviations: FADE = functional activity deviation during encoding, SAME = similarities of activations during memory encoding, A-prime = memory performance, BMI = body-mass index, MMSE = mini-mental state examination^45,44^, NPT = neuropsychological testing^46^, PACC5 = preclinical Alzheimer’s cognitive composite including the category fluency measure^47^. Significance: * p < 0.05, Bonferroni-corrected for ** number of independent variables (16) or *** number of variables and number of scores (16 x 4).

### Effects of ApoE genotype in AD relatives

Before assessing effects of ApoE genotype on fMRI scores, we investigated the distribution of ApoE genotypes within each diagnostic group. We computed chi-squared goodness-of-fit tests comparing the actual occurrences of genotypes to expected frequencies obtained from a comparable population^60^.

Individuals with MCI or AD differed significantly from the population distribution with higher frequencies of ε3/ε4 and ε4/ε4 (Figure S5 and Table 3), compatible with the higher frequency of the ε4 allele in AD.

A between-subjects ANOVA on the fMRI scores with diagnostic group (HC, SCD, MCI, AD, AD-rel) and ApoE genotype (ε3 homozygotes vs. ε4 carriers) as fixed factors, yielded a significant main effect of ApoE for all scores except the novelty-FADE score, but not when correcting for multiple comparisons (novelty-FADE: F_1,393_ = 1.11, p = 0.293; novelty-SAME: F_1,393_ = 5.14, p = 0.024; memory-FADE: F_1,393_ = 6.54, p = 0.011; memory-SAME: F_1,393_ = 4.24, p = 0.040). When calculating post-hoc tests comparing the scores between ApoE ε4 carriers and ε3 homozygotes in each diagnostic group, we found significant differences among the AD relatives (novelty-FADE: t_41_ = −2.56, p = 0.014; novelty-SAME: t_41_ = 2.45, p = 0.019; memory-FADE: t_41_ = −2.20, p = 0.034; memory-SAME: t_41_ = 1.48, p = 0.146), but not in other groups (all p > 0.058; Figure 4). Thus, the increased genetic risk in AD relatives was also reflected by FADE and SAME scores.

**Figure 4.**
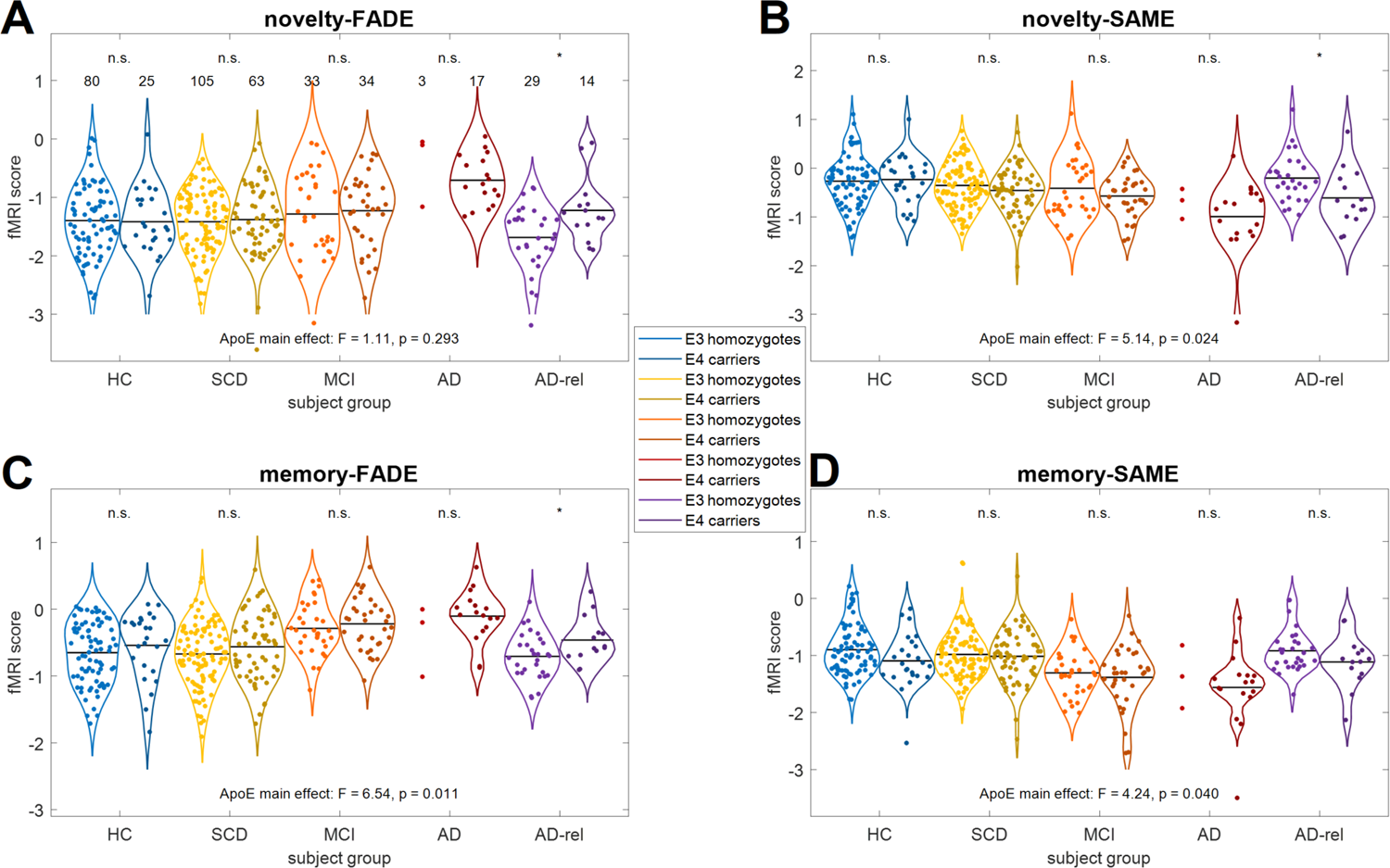
FADE and SAME scores by diagnostic group and ApoE genotype. Single-value fMRI scores are shown for **(A)** the FADE score and **(B)** the SAME score computed from the novelty contrast as well as **(C)** the FADE score and **(D)** the SAME score computed from the memory contrast. The layout follows that of Figure 2. Sample sizes are given in the upper-left panel. Horizontal bars correspond to group-wise means. Violin plots and group means are not shown for sample sizes N ≤ 5. Markers on top of the x-axis denote a two-sample t-test between ε4 carriers (ApoE variants ε2/ε4, ε3/ε4 and ε4/ε4) and ε3 homozygotes (ApoE genotype ε3/ε3; n.s. = not significant; * p < 0.05).

### Effects of amyloid positivity in SCD and AD relatives

Finally, we examined a potential association of the scores with amyloid positivity, defined by the CSF Aβ-42/40 ratio (value ≤ 0.08 considered as A+; according to ^48^). Initial omnibus between-subjects ANOVAs with diagnostic group (HC, SCD, MCI, AD, AD-rel) and amyloid positivity (A+, A–) as fixed factors revealed a main effect of Amyloid for all scores, except for the memory-SAME score, after correcting for multiple comparisons (novelty-FADE: F_1,214_ = 12.46, p < 0.001; novelty-SAME: F_1,214_ = 10.59, p = 0.001; memory-FADE: F_1,214_ = 7.13, p = 0.008; memory-SAME: F_1,214_ = 4.25, p = 0.040). When calculating post-hoc t-tests comparing participants by Amyloid status within each diagnostic group, we found that these effects were driven by higher FADE scores and lower SAME scores in A+ participants across all diagnostic groups. These differences were significant in individuals with SCD (novelty-FADE: t_90_ = 2.57, p = 0.012; novelty-SAME: t_90_ = −2.52, p = 0.013; memory-SAME: t_90_ = −2.05, p = 0.044) and AD relatives (novelty-SAME: t_22_ = −2.70, p = 0.013), but not in the other groups (all other p > 0.061; Figure 5). Thus, our scores, especially the novelty-based scores, were indeed sensitive to amyloid positivity. However, given the small sample size in some subgroups (e.g., AD patients with A–, AD relatives with A+), those findings must be considered preliminary.

**Figure 5.**
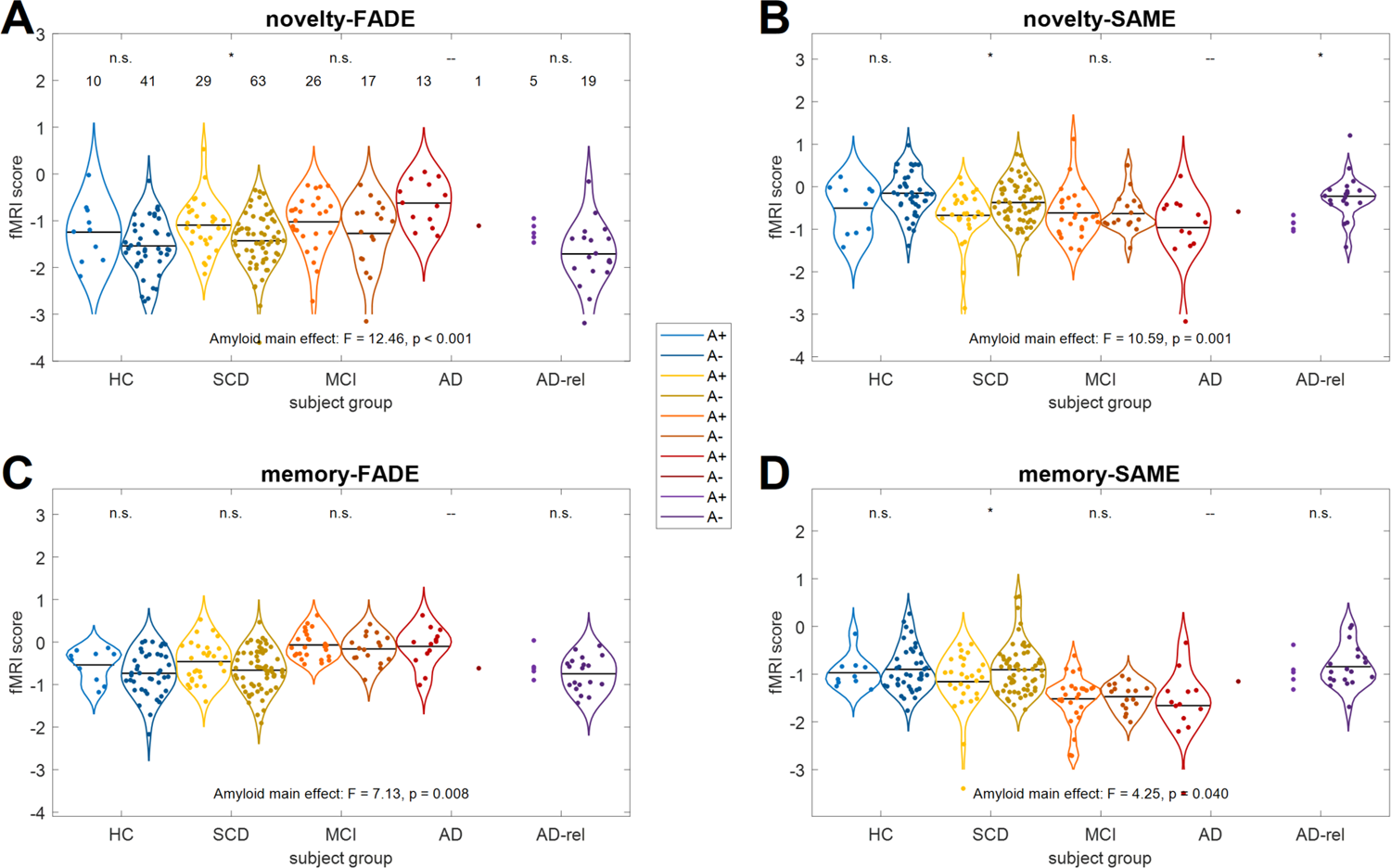
FADE and SAME scores by diagnostic group and amyloid positivity. Single-value fMRI scores are shown for **(A)** the FADE score and **(B)** the SAME score computed from the novelty contrast as well as **(C)** the FADE score and **(D)** the SAME score computed from the memory contrast. The layout follows that of Figure 2. Sample sizes are given in the upper-left panel. Horizontal bars correspond to group-wise means. Violin plots and group means are not shown for sample sizes N ≤ 5. Markers on top of the x-axis denote a two-sample t-test between Amyloid-positive (A+: Aβ 42/40 ≤ 0.08) and Amyloid-negative (A–: Aβ 42/40 > 0.08) individuals (n.s. = not significant; * p < 0.05).

### Predictive utility of FADE and SAME scores

When using all four scores for SVM classification, diagnostic group could be predicted with above-chance classification accuracy for several partitions, such as distinguishing all five groups (all participants; balanced accuracy, BA = 31.17%, confidence interval, CI = [23.54%, 39.12%]) or the clinical groups (SCD, MCI, AD; BA = 49.27%, CI = [38.21%, 60.05%]), but also MCI and AD from healthy controls (MCI vs. HC: BA = 68.94%, CI = [61.99%, 75.09%]; AD vs. HC: BA = 78.03%, CI = [64.97%, 87.72%]).

In AD relatives, ApoE genotype could be predicted above chance (BA = 68.44%, CI = [51.37%, 82.39%]). The same was true for classification of Amyloid status in individuals with SCD, but the confidence interval did not exclude chance level due to small sample sizes (BA = 55.65%, CI = [43.88%, 66.52%]; for details, see Supplementary Table S4).

## Discussion

We have explored the utility of single-value scores derived from memory-related fMRI contrast maps as potential biomarkers across the AD risk spectrum (SCD, MCI, and AD, plus AD-rel). We could replicate and extend earlier findings on the neurocognitive underpinnings of FADE and SAME scores in healthy older adults^29,30^ and identified several characteristic associations of the scores with neurobiological markers of AD risk. Among healthy older adults, we could largely replicate our previous findings in voxel-wise fMRI data analysis (Figures 1C/D and S1). Single-value scores also showed similar associations with neurocognitive measures and nuisance variables (Figures 2, 3, 4, S6, and S7; Table 2; Supplementary Results).

### FADE and SAME scores across the AD risk continuum

In line with our hypothesis, the fMRI scores show a continuous increase (FADE scores) or decrease (SAME scores) across AD risk spectrum stages (Figure 2). In the SCD group, we observed nominally higher FADE and lower SAME scores compared to HC, but the overall pattern was largely preserved. Individuals with MCI, on the other hand, showed markedly higher FADE scores and lower SAME scores for the memory contrast, whereas the novelty-based scores showed only gradual differences to those from the SCD group (similar in magnitude as between the HC and SCD groups). In the AD group, we additionally observed markedly altered scores for the novelty contrast, which distinguished them from the MCI group. These findings suggest that subsequent memory effects, and thus the FADE and SAME scores computed from the memory contrast, might be more sensitive to small deviations from typical memory processing, as they also reflect encoding success.

While these results are generally compatible with the notion that SCD and MCI can be considered intermediate stages between healthy brain aging and manifest AD, they additionally suggest qualitative differences with a substantial disruption of memory encoding-related brain activity distinguishing MCI from SCD and an additional (i.e., more substantial) impairment of novelty processing marking the transition from MCI to AD. Accelerated forgetting, resulting in impaired long-term recall is impaired already early in the AD continuum. Specifically, recall after prolonged retention intervals (e.g., several days) can be affected at pre-MCI stages, whereas MCI is associated with impaired recall after intermediate retention intervals^61,62^, such as the 70 min employed here, thereby allowing for a differentiation between individuals with MCI versus SCD. On the other hand, the additional effect on the novelty-based scores in participants with manifest AD may be best explained by a broader deficit present at the initial encoding stage already^61,62^. Within this framework, future studies should further explore the relationship between FADE and SAME scores and retrieval after prolonged retention intervals in individuals with SCD.

### Brain activity patterns underlying FADE and SAME scores

When comparing voxel-wise fMRI contrasts across diagnostic groups, we found that differences in scores could be attributed to both reduced temporo-parieto-occipital memory network activations and reduced DMN deactivations (Figure 1). They thus mirror previously described activation differences between healthy older adults and individuals with MCI or AD^31–34,24^. Qualitatively, these patterns are similar to memory-related fMRI activation differences between healthy young and older adults^25,29,51^. One interpretation of the observed pattern would therefore be that progressive deterioration of memory-related brain activity across the AD risk spectrum might reflect accelerated neurocognitive aging.

Notably, individuals with SCD exhibited largely preserved temporo-parietal memory network activations during novelty processing and successful encoding, but reduced novelty-related deactivations of DMN structures like the precuneus (Figure 1), replicating previous results based on a different first-level GLM^24^. This observation is compatible with earlier findings suggesting that age-related reduced deactivations of DMN structures are associated with lower memory performance^25^ and with the notion that reduced inhibitory activity may constitute an early mechanism of neurocognitive aging^63,64^.

In the more severely affected diagnostic groups (i.e., MCI and AD), we additionally observed reduced activations of the MTL and parieto-occipital memory network structures (Figures 1 and S1). In the novelty contrast, these were primarily observed in AD patients, whereas both the MCI and the AD group exhibited reduced temporo-parieto-occipital network activity in the memory contrast, reflecting the pattern of FADE and SAME scores. Using Bayesian model selection of first-level fMRI models, we found that in both groups, a memory-invariant model was favored over even the most parsimonious subsequent memory models^38^. Additionally, the AD group also showed a substantially lower number of voxels favoring a novelty model over a purely perceptual model not considering novelty. Therefore, a straightforward explanation for the higher FADE and lower SAME scores in the MCI and AD groups may be that the memory contrasts and – in the case of the AD group, also the novelty contrasts – underlying the scores might exhibit a lower signal-to-noise ratio resulting from a suboptimal model fit in these diagnostic groups.

### FADE and SAME scores as indices of neurocognitive aging

Across the cohort, FADE and SAME scores correlated with neuropsychological measures like MMSE, NPT and PACC5, after controlling for diagnostic group (Figure 3). This pattern is in line with previous observations that the scores reflect indices of neurocognitive age differences^22,29,30^. A previous evaluation of the FADE and SAME scores in healthy older adults has suggested that all scores correlate with delayed episodic recall performance, and memory-based scores additionally correlate with more global measures of cognition^30^.

While we previously found no correlation between memory performance in the fMRI task and the FADE score derived from the novelty contrast^29,30^, this correlation was significant in the present study, possibly due to a larger sample size. We could nevertheless replicate the observation that memory performance in the fMRI task showed a stronger correlation with the scores computed from the memory contrast as compared to the novelty contrast^29^. Correlations with independent neuropsychological indices (NPT global, PACC5 score) were similar in magnitude across the four scores, albeit nominally stronger for the novelty-based scores, tentatively suggesting a potentially higher prognostic value with respect to prediction of cognitive functioning in individuals at risk for AD. That said, computing the scores from the memory contrast may nevertheless be beneficial for differentiating individuals with SCD from individuals with MCI (Section “FADE and SAME scores across the AD risk continuum“). Furthermore, particularly the memory-SAME score may be suitable for the prediction of individual differences of cognitive performance in healthy older adults^39,30,65^.

### Single-value scores, amyloid status, and genetic risk

While the differential patterns of FADE and SAME scores observed here (Figure 2) allow for a separation of individuals with SCD, MCI, and AD, their diagnostic value for differentiating individuals with SCD from healthy controls is less clear. Likewise, scores in healthy relatives of patients with AD were essentially indistinguishable from those of the HC group. However, these groups exhibited specific associations between the scores and markers of Alzheimer’s pathology (Aβ-42/40 ratio) and genetic risk (ApoE ε4 allele carriage).

Among all participants with available CSF samples, novelty-based FADE and SAME scores differed as a function of Aβ-42/40 ratio (Figure 3). When testing for effects of amyloid positivity separately in each group, the effect was only significant in the SCD (novelty-FADE and SAME scores) and AD-rel groups (novelty-SAME score; Figure 5). This observation opens a potential perspective for the scores as diagnostic or prognostic markers of AD risk in SCD. Individuals with SCD typically report memory problems, despite objectively normal or only mildly impaired neuropsychological test performance^12,13^, and minor neuropsychological deficits in SCD have been linked to reduced Aβ-42/40 ratios and increased p-tau181 levels in CSF^46^. Amyloid positivity in SCD has recently been associated with subsequent clinical progression to MCI^48^ and with lower hippocampal volumes^66^. Therefore, FADE and SAME scores – and perhaps particularly the novelty-SAME score – may constitute novel non-invasive predictors for the progression to MCI in individuals with SCD.

A similar pattern was found in AD relatives whose FADE and SAME scores did, on average not differ from those of HC. Unlike previous studies of neuropsychological performance in healthy relatives of patients with AD (for a review, see ^67^), we additionally found no performance difference between healthy relatives and control participants (see Fig. 1 in ^38^). However, unlike HC and similar to individuals with SCD, healthy relatives exhibited a significant effect of amyloid positivity on the novelty-SAME score (Figure 5). This is in line with the observation that, in the same cohort, amyloid positivity has been associated with higher subjective cognitive decline in the relatives^46^. Additionally, AD relatives were the only group in which we found an association of the scores with ApoE genotype (Figure 4). This suggests that indices of subtle cognitive impairment in relatives of patients with AD (i.e., higher FADE and lower SAME scores) reflect, at least in part, genetic risk and is compatible with previously reported synergistic effects of ApoE ε4 carriage and AD family history on brain amyloid deposition^68^. Note that relatives carrying the ApoE ε4 allele have previously been shown to display lower performance in cognitive tests^67^. While ApoE ε4 carriage, and particularly ε4 homozygosity, is the strongest risk factor for sporadic (late-onset) AD, future studies should further assess the role of polygenic risk on fMRI-based scores and their trajectories in relatives of AD patients.

### Limitations and directions for future research

One limitation of our study is that the BOLD signal underlying the fMRI activation patterns and thus FADE and SAME scores is an indirect measure of neural activity and profoundly influenced by vascular and metabolic factors. While dynamic cerebral autoregulation, a key mechanism of regulating cerebral blood flow, is largely preserved, at least macroscopically, in MCI and AD^69^, small-vessel disease like Amyloid angiopathy is commonly associated with AD and can impair neurovascular coupling^70^, which may in turn contribute to a blunted BOLD signal, particularly in MCI or AD. On the other hand, even if the pattern of FADE and SAME scores in the MCI and AD groups can, at least partly, be attributed to vascular or metabolic differences, this should not necessarily affect their potential diagnostic value. However, caution is warranted with respect to the interpretation of underlying neural mechanisms.

Another limitation concerns the composition of the sample, as participant groups significantly differed regarding age range, gender ratio, acquisition site, ApoE genotype, CSF biomarkers and neuropsychological measures (Table 1). While some of these imbalances directly result from the study design, reflecting expected differences in neuropsychological scores and fluid biomarkers, other variables like age or gender constitute potential confounds. Here, we aimed to statistically control for such factors while maximizing the sample size to increase statistical power. It must be noted, though, that, for example, gender effects may be worthwhile to investigate in more detail^71,72^. On the other hand, the sample was ethnically and socio-demographically rather homogenous, most likely owing to our recruiting strategy via memory clinics and newspaper advertisements. Further studies should assess the generalizability of our findings to individuals from different ethnic and cultural backgrounds^73,74^.

Furthermore, while SVM classifications allowed us to explore the predictive utility of the scores to some extent, a longitudinal study is needed to assess whether the scores actually bear a prognostic value in individuals at risk for AD.

Another limitation is that the FADE and SAME scores are inherently linked to a reference cohort. We have previously shown their robustness with respect to different reference samples of young adults, although smaller samples were associated with steeper slopes and non-zero intercepts^29^. A similar relationship was found when using a sample of healthy older adults of similar size as reference (Supplementary Methods, Results, and Figure S4). Importantly, the relationship between the scores based on different reference samples was essentially linear, thus affecting their absolute value, but not their distribution in the study population (see Supplementary Discussion). Ultimately, the strongest evidence for the robustness of the scores would, in our view, be a proof of test-retest reliability, possibly moderated by cognitive decline (i.e., scores of subsequent decliners being less stable over time than those of individuals with longitudinally preserved cognitive function) and/or Amyloid status. This will be addressed in future work.

## Conclusions

We have demonstrated a use case for reductionist single-value scores, computed from whole-brain fMRI contrast maps, across the trajectories of the AD risk spectrum in a cross-sectional design. FADE and SAME scores vary as a function of disease status group (i.e., MCI vs. AD), whereas in individuals with moderately elevated risk (i.e., SCD and AD-rel), the scores distinguish individuals with and without additional risk factors (i.e., Aβ 42/40 ratio, ApoE genotype). Our results demonstrate the potential utility of FADE and SAME scores as fMRI-based biomarkers for neurocognitive functioning in individuals at risk for AD, but longitudinal studies are needed to evaluate a potential prognostic use.

## Data availability

Data from the DELCODE study are available via individual data sharing agreements with the DELCODE study board (for more information, see https://www.dzne.de/en/research/studies/clinical-studies/delcode/). The code used for computing the FADE and SAME scores has been published previously^29^ and is available via GitHub (https://github.com/JoramSoch/DELCODE_SAME).

## Acknowledgements

We would like to thank all the participants in the DELCODE study and all the technical, medical and psychological staff for making this study possible. Special thanks go to the Max Delbrück Center for Molecular Medicine (MDC) within the Helmholtz Association, the Center for Cognitive Neuroscience Berlin (CCNB) at the Free University of Berlin, the Bernstein Center for Computational Neuroscience (BCCN) Berlin, the MR research core facility of the University Medical Center Göttingen (UMG) and the MR research center of the University Hospital Tübingen (UKT).

## Supplementary material

Supplementary material is available at *Brain* online.

## Appendix

### Parametric modulator reflecting encoding success

In the voxel-wise GLM for first-level fMRI analysis, values for the parametric modulator (PM) regressor were given by

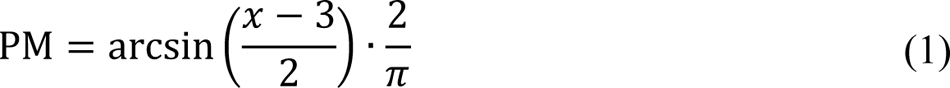

where *x* ∈ {1, 2, 3, 4, 5} is the later subsequent memory report, such that the transformation ensures that – 1 ≤ PM ≤ +1.

The use of the arcsine function in the transformation causes definitely remembered or forgotten items (1, 5) to be weighted stronger relative to probably remembered or forgotten items (2, 4) than when using a linear mapping.

### Calculation of single-value fMRI scores

Let *J*_−_ and *J*_+_ be the sets of voxels showing a negative effect or a positive effect, respectively, on a particular contrast in young subjects at an *a priori* defined significance level (p < 0.05, FWE-corrected, extent threshold k = 10), and let *t*_ij_ be the t-value of the *i*-th older subject in the *j*-th voxel on the same contrast. Then, the FADE score of this subject is given by

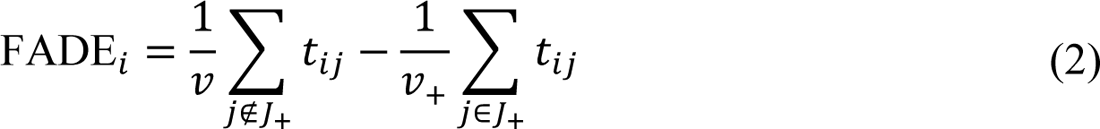

where *v*_+_ and *v* is the number of voxels inside and outside *J*_+_, respectively.

Alternatively, let β^^^_j_ be the average estimate on a particular contrast in young subjects, let σ^^^_j_ be the standard deviation of young subjects on this contrast at the *j*-th voxel and let γ^^^_ij_ be the contrast estimate of the *i*-th older subject at the *j*-th voxel. Then, the SAME score of this subject is the sum of averaged reduced activations in *J*_+_and averaged reduced deactivations in *J*_-_

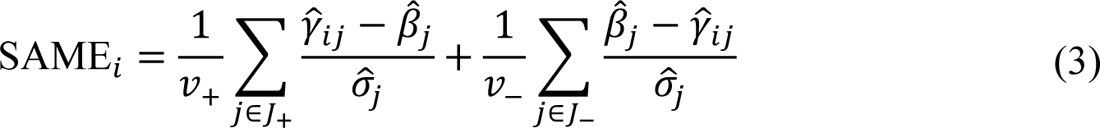

where *v*_+_ and *v*_-_ are the numbers of voxels in *J*_+_ and *J*_-_, respectively.

