## Supplementary Text, Figures and Tables for "Single-value brain activity scores reflect both severity and risk across the Alzheimer’s continuum"

### Supplementary Methods

##### Cross-validated vs. out-of-sample calculation of FADE and SAME scores

In the original study, because we intended to also determine FADE and SAME scores for young subjects, young and older adults were partitioned into cross-validation (CV) groups and each subject's scores were determined using only young subjects from the respective other CV group as reference (see Soch et al., 2021b, Section 2.9 and Table S3). The original study also included a comparison (see Soch et al., 2021b, Figure S7) of either using all young subjects (contrary to the CV scheme) or just half of those subjects (equivalent to the CV scheme) as the reference group, culminating in the conclusion that "when focusing on neural processes underlying age-related memory decline […], scores should be calculated based on all available young subjects" (Soch et al., 2021b, suppl. p. 5).

In accordance with this, we therefore extracted FADE and SAME scores for the DELCODE sample with reference to the entire group of young healthy subjects (N = 106; see Soch et al., 2021b, Table 1). To render these FADE and SAME scores comparable with those from the original study, the latter were re-calculated also using the whole set of young subjects as the reference sample. This has the consequence that fMRI scores for young subjects (e.g., as reported in Figure 2) are partly circular[1] and should not be interpreted. However, as the focus of the present study is not on young subjects, but rather on the comparison of the different patient groups (SCD, MCI, AD) with healthy older adults (HC), young subjects are displayed for purely illustrative purposes.

##### Stability of FADE and SAME scores as a function of reference sample

To assess the stability of FADE and SAME scores with respect to the utilized reference sample, we compared scores computed with respect to young subjects in the original study (reference sample "young AiA"; Soch et al., 2021b) to scores computed based on an independent sample of young adults (reference sample "yFADE"; Assmann et al., 2020). That cohorts were administered the same fMRI paradigm, with minor differences in trial timings, data acquisition, and preprocessing (cf. Table S1 in Soch et al., 2021a). To assess how FADE and SAME scores change when using a reference sample not consisting of young adults, we also compared the original scores to scores computed based on the older subjects in the original study (reference

[1] Another consequence of this is that the SAME scores for young subjects have mean zero (see Figure 4B/D). This follows from the construction of the SAME score (see Soch et al., 2021, Section 3.3): Since the fMRI activations of young adults are on average not different from the average fMRI activations in young adults, they are exactly matching the reference sample on average (see Soch et al., 2021, last eq. on p. 6).

sample "old AiA"; Soch et al., 2021b).

Relationships between re-computed scores with original scores were fit as regression lines of the form y = mx + n for the two comparisons "yFADE vs. young AiA" and "old AiA vs. young AiA". Regression coefficients m and n were compared between fits via paired t-tests to assess which scores further deviated from the identity line when compared against the original scores.

**Analysis of FADE and SAME scores as a function of educational status**

In the original study, in lack of a more precise measure such as educational years, we used the "Mehrfachwahl-Wortschatz-Intelligenztest" (MWT-B; Lehrl, 1999, 2005), a vocabulary-based screening of verbal intelligence, and the presence of "Abitur", i.e. the German equivalent of a high-school diploma, as surrogate measures for educational background. While the MWT-B was not administered to DELCODE subjects, information about Abitur is available[2], such that the corresponding analysis could be run in the present study.

**Analysis of FADE and SAME scores as a function of chronological age**

In the original study, we visualized FADE and SAME scores as a continuous function of chronological age which highlighted that fMRI scores are dominated by age group effects rather than effects of age within age groups (see Soch et al., 2021b, Figure S5).

Here, we analyzed single-value scores in the same way, by plotting FADE and SAME scores from all subjects along with the smooth mean and smooth variance as a function of age, separated by age group (HC, SCD, MCI, AD, AD-rel) and using a sliding window of 32 years for the whole age range from 18 to 90 (see Figure S6; identical to original study) and a sliding window of 16 years for the older age range from 60 to 90 (see Figure S7).

[2] For the present analysis, the entries "Abitur" (= high-school diploma) and "Fachabitur" (= technical diploma) were regarded as "with Abitur" and all other graduations from school were considered "without Abitur".

### Supplementary Results

**There is an interaction of score type and diagnostic group on fMRI scores**

In order to compare the modulation of the different scores by Alzheimer's disease state, we performed a two-way mixed ANOVA with (i) diagnosis group (HC, SCD, MCI, AD, AD-rel) as between-subject factor and (ii) type of fMRI contrast (novelty, memory) as within-subject factor, separately for FADE scores and SAME scores.

There was a significant main effect of contrast and a significant interaction of diagnosis and contrast for both types of scores (see Table S3). Between-group score differences are nominally larger for the SAME scores (novelty-FADE: $\Psi = 0.72$; novelty-SAME: $\Psi = 0.84$; memory-FADE: $\Psi = 0.88$; memory-SAME: $\Psi = 0.99$), estimated as root-mean-square standardized effect $\Psi$ (without AD relatives, to capture changes across the AD risk spectrum), the multi-group analogue to Cohen's d (Steiger, 2004). This means that, although both scores show significant effects of diagnosis (see Table 2), SAME scores show nominally larger differences between AD risk states (see Figure 2).

**Single-value scores are stable for young subjects, but deviate for older subjects**

For all score types (novelty/memory x FADE/SAME), scores computed based on young AiA subjects were highly correlated to their respective scores calculated using the yFADE sample (all $r > 0.96$, all $p < 0.001$; see Figure S3) and also highly correlated to their respective scores calculated based on old AiA subjects (all $r > 0.96$, all $p < 0.001$; see Figure S4).

However, regression coefficients were close to the identity line for the yFADE comparison (all $m \approx 1$, all $n \approx 0$) and more apart from those values for the old AiA comparison (typically $m > 1$ and $n > 0$). This was particularly true for the SAME scores, but not for the novelty-FADE score (see Table S5). The intercept term not significantly differing for the novelty-FADE scores can be explained by the fact that young and older subjects also did not differ with respect to this score in the original study (see Soch et al., 2021b, Table 2, Figures 2 and S5).

**There are no robust effects of educational status on fMRI scores**

When performing a between-subject ANOVA with diagnostic group (HC, SCD, MCI, AD, AD-rel) and educational status (with vs. without Abitur), the main effect of Abitur was nominally significant only for the memory-SAME score, but not when correcting for multiple comparisons (novelty-FADE: $F_{1,458} = 0.10$, $p = 0.749$; novelty-SAME: $F_{1,458} = 1.84$, $p = 0.175$; memory-FADE: $F_{1,458} = 1.99$, $p = 0.159$; memory-SAME: $F_{1,458} = 5.20$, $p = 0.023$). There were no significant interactions of Abitur and diagnostic group (all $p > 0.083$).

When calculating post-hoc tests comparing the with/without Abitur groups in each diagnostic group, we found that this effect was supported by significant differences of SAME scores within individuals with SCD (novelty-SAME: $t_{197} = 2.43$, $p = 0.016$; memory-SAME: $t_{197} = 3.10$, $p = 0.002$), but not in any other group (see Figure S5). This coincides with findings from the original study, where also almost no effects of MWT-B or Abitur on FADE and SAME scores were observed (see Soch et al., 2021b, Figure S4).

**There are no robust associations of fMRI scores with chronological age**

In the original study, we visualized FADE and SAME scores as a continuous function of chronological age to show that, although there are strong age group effects, the fMRI scores are largely age-independent within age groups (see Figure S6). Similarly, when analyzing data from the DELCODE study, we found that differences in old age were largely driven by diagnostic group rather than by chronological age, as indicated by nearly flat trajectories of FADE and SAME scores as a function of age in all diagnostic groups (see Figure S7).

### Supplementary Discussion

**Single-value scores and their associations replicate in healthy older adults**

In healthy older adults, we could largely replicate our previous findings. In voxel-wise fMRI data analysis, we found novelty processing and subsequent memory to engage overlapping temporo-parieto-occipital networks, accompanied by default mode network (DMN) deactivations (see Figures 1C/D and S1; see Soch et al., 2021b, Fig. 2), also replicating earlier studies with other stimulus types and encoding tasks (Maillet & Rajah, 2014; Kim, 2011). Please note that, despite their anatomical overlap, novelty processing and successful encoding both contributed to fMRI signal variance explanation within the memory network in the HC, SCD, and AD-rel groups, albeit not in the MCI and AD groups (Soch et al., 2023).

Compatibly, we found that single-value scores from the HC group were statistically indistinguishable from those of healthy older adults in the preceding study (see Figures 2 and S7). Importantly, we could also replicate the core associations (or lack thereof) with demographic variables and confounding factors (Soch et al., 2021b):

1. Acquisition site and scanner had no effect on any of the scores (see Table 2).
2. Gender had negligible effects on the scores and showed no interactions (see Figure S2).
3. FADE and SAME scores showed high reliability when computed with different reference samples of young adults (see Figure S3).

As in the original study, SAME scores were negatively correlated with age and positively correlated with memory performance; FADE scores showed the reverse pattern, reflecting their construction. Unlike in the original study, correlations of memory performance with the FADE score computed from the novelty contrast were also significant (see Figure 3). Finally, in line with the original, we found no significant associations of the scores with either ApoE genotype or educational status in the HC group (see Figures 4 and S6). In summary, the overall replication of patterns of association points to the robustness and reliability as a prerequisite for the scores' potential use as biomarkers.

**Potential influences of different reference samples**

As the scores are inherently dependent on the activation patterns in the reference sample, we conducted control analyses during both the present study and as part of our initial description of the FADE and SAME scores (Soch et al., 2021b). In all of these analyses, we did not change the contrasts on which the calculation of scores was based, as fMRI contrasts outside the temporo-parieto-occipital memory networks would trivially not affect the scores.[3] Instead, the entire reference sample on which prototypical activations were based was varied, while always computing the scores based on the same contrasts. In our initial study, we have investigated how the number of subjects in the young reference sample impacts estimation of FADE and SAME scores. When using smaller reference samples (N = 53, i.e., half the reference sample), the scores were highly correlated with those based on the full reference sample, although the slope of the regression line was steeper than the identity line, most likely due to a smaller extent of activation clusters and larger standard errors in the activation maps derived from the smaller samples (see Soch et al., 2021, Fig. S7).

In the present study, we additionally assessed the influence of the reference sample's age group. When correlating FADE and SAME scores with those based on an independent young cohort of similar size (N ≈ 100), correlation coefficients were close to one and regression lines were close to the identity line (see Figure S3). This pattern changed when FADE and SAME scores were based on a non-young cohort, i.e., older participants from the cohort of the original study (Soch et al., 2021b). Similar to the scores based on the independent sample of young adults, scores based on an older reference sample correlated strongly with the scores computed based on the young cohort (see Figure S4). Notably, similarly to the scores based on smaller samples of young adults (Soch et al., 2021b), the intercept and slope of the corresponding regression lines differed significantly (see Table S5). Specifically, the slope was significantly steeper for all scores except for the novelty-FADE score, and the intercept is significantly positive for the SAME scores and significantly negative for the memory-FADE score. This exception of the novelty-FADE score is compatible with the earlier finding that the novelty-FADE score does not differ significantly between age groups (see Soch et al., 2021, Fig. 3 and Fig. S5). The steeper slopes and non-zero intercepts most likely reflect the phenomenon that individuals with very high SAME scores (or low memory-FADE scores) display "above-average" scores when using a non-young reference sample.

Importantly, all changes to the scores with respect to different reference samples were essentially very close to linear transformations, as evident from the very high – and highly significant – correlation coefficients and a narrow distribution around the regression lines. Therefore, even though size and age group of the reference sample can influence the values of the scores, their influence on the distribution of the scores within a given population is minor and likely negligible.

---

[3] An exception may be the deactivations in DMN structures that contribute to the SAME scores, provided that individual differences in DMN deactivations are task-independent (Kizilirmak et al., 2023).

#### Supplementary Tables

| Analysis | Description | original study | present paper | this supplement |
|---|---|---|---|---|
| 1 | fMRI novelty and memory effect as a function of participant group | Figure 2 | Figure 1 | Figure S1 |
| 2 | FADE/SAME scores as a function of scanner (here: site), gender and group | Table 2 | – | Figure S2 |
| 3 | FADE/SAME scores as a function of age group (here: diagnosis) and score type | Table 3<br>Figure 3 | Table 2<br>Figure 2 | Table S3<br>Table S4 |
| 4 | correlation of FADE/SAME scores with other indices of cognitive aging | Figure 4<br>Figure S6 | Figure 3 | – |
| 5 | stability of FADE/SAME scores for young adults across different studies | Figure 5 | – | – |
| 6 | stability of FADE/SAME scores for older adults across reference samples | Figure 6 | – | Figure S3<br>Figure S4 |
| 7 | distribution of ApoE genotype for each age group (here: diagnosis) | Figure S3A | Table 3 | Figure S5 |
| 8 | FADE/SAME scores as a function of participant group and ApoE genotype | Figure S3B | Figure 4 | – |
| 9 | FADE/SAME scores as a function of participant group and educational status | Figure S4B | – | Figure S6 |
| 10 | relationship of FADE/SAME scores with chronological age | Figure S5 | – | Figure S7<br>Figure S8 |
| 11 | extraction of FADE/SAME scores from all vs. half of young subjects | Figure S7 | – | – |
| 12 | FADE/SAME scores as a function of participant group and Amyloid positivity | – | Figure 5 | – |

**Table S1.** *Index of statistical analyses for fMRI scores.* This table lists group-level fMRI analyses conducted in the original study and replicated for the present paper. Note that the factors "fMRI scanner" and "age group" in the original study were conceptually replaced with "acquisition site" and "diagnostic group" for the present paper. The last three columns list where to find results in the original paper, in the main manuscript and in this supplement.

| Step of data acquisition/processing | Description in Soch et al., 2021 |
|---|---|
| experimental paradigm | see Section 2.2 |
| fMRI data acquisition | see Section 2.3 |
| fMRI data preprocessing | see Section 2.4 |
| general linear modelling | see Section 2.5 |
| single-value fMRI scores | see Sections 2.6/2.7 and Figure 1 |
| extraction of fMRI scores | see Section 2.8 and Table S3 |
| group-level statistical analyses | see Section 2.9 |

**Table S2.** *Reference for data acquisition and processing.* Steps of data acquisition and processing are summarized in Sections 2.2 to 2.6 and 2.9 of the main manuscript. Details can be found in the referenced sections of the original publication (right column).

| | FADE scores | SAME scores |
|---|---|---|
| main effect of diagnosis | $F_{4,463} = 17.60$, $p < 0.001$ | $F_{4,463} = 19.24$, $p < 0.001$ |
| main effect of contrast | $F_{4,463} = 877.65$, $p < 0.001$ | $F_{4,463} = 1516.41$, $p < 0.001$ |
| interaction of diagnosis and contrast | $F_{4,463} = 18.78$, $p < 0.001$ | $F_{4,463} = 19.80$, $p < 0.001$ |

**Table S3.** *Effects of diagnosis group and fMRI contrast on single-value scores.* Results from two-way ANOVAs with diagnostic group (HC, SCD, MCI, AD, AD-rel) and fMRI contrast (novelty, memory) as factors for FADE scores and SAME scores. This table corresponds to Table 3 from the original publication.

| Classification analysis | sample sizes | chance level | balanced accuracy | 90% confi-dence interval | lower CI above CL? |
|---|---|---|---|---|---|
| 5 groups (HC, SCD, MCI, AD, AD-rel) | 128, 199, 74, 21, 46 | 0.20 | 0.3117 | [0.2354, 0.3912] | yes |
| 4 groups (HC, SCD, MCI, AD) | 128, 199, 74, 21 | 0.25 | 0.3781 | [0.2875, 0.4700] | yes |
| 3 groups (HC, MCI, AD) | 128, 74, 21 | 0.33 | 0.5133 | [0.4016, 0.6200] | yes |
| 3 groups (SCD, MCI, AD) | 199, 74, 21 | 0.33 | 0.4927 | [0.3821, 0.6005] | yes |
| HC vs. SCD | 128, 199 | 0.50 | 0.5427 | [0.4894, 0.5952] | no |
| HC vs. MCI | 128, 74 | 0.50 | 0.6894 | [0.6199, 0.7509] | yes |
| HC vs. AD | 128, 21 | 0.50 | 0.7803 | [0.6497, 0.8772] | yes |
| HC vs. SCD | 128, 199 | 0.50 | 0.5427 | [0.4894, 0.5952] | no |
| SCD vs. MCI | 199, 74 | 0.50 | 0.6378 | [0.5668, 0.7024] | yes |
| MCI vs. AD | 74, 21 | 0.50 | 0.6072 | [0.4645, 0.7283] | no |
| ApoE: ε3 homozygotes vs. ε4 carriers (controlling for group) | 250, 153 | 0.50 | 0.5430 | [0.4940, 0.5906] | no |
| ApoE: ε3 homozygotes vs. ε4 carriers (AD-rel only) | 29, 14 | 0.50 | 0.6844 | [0.5137, 0.8239] | yes |
| Amyloid: pos. vs. neg. (controlling for group) | 83, 141 | 0.50 | 0.6019 | [0.5338, 0.6639] | yes |
| Amyloid: pos. vs. neg. (SCD only) | 29, 63 | 0.50 | 0.5565 | [0.4388, 0.6652] | no |

**Table S4.** *Results from support vector classifications based on single-value scores.* Sample sizes, chance levels, balanced accuracies (Brodersen et al., 2010) and 90% confidence intervals are given for different classification analyses. The first column details which distinct participant groups were used for classification. The last column specifies whether the confidence interval excludes the chance level, indicating significant above-chance classification accuracy. Balanced accuracies and confidence intervals were obtained as averages from 1000 subsamples where each subsample was drawn, such that all classes have the same number of samples, equal to the number of samples in the smallest class.

| Score | Group | yFADE vs. young AiA (y = mx + n) | old AiA vs. young AiA (y = mx + n) | Statistics |
|---|---|---|---|---|
| novelty-FADE | HC | y = 1.10 x – 0.01 | y = 0.95 x + 0.01 | m: yFADE > AiA old; |
| | SCD | y = 1.11 x – 0.00 | y = 0.94 x – 0.01 | t = 29.14, p < 0.001 |
| | MCI | y = 1.09 x – 0.01 | y = 0.94 x – 0.00 | n: not significant; |
| | AD | y = 1.07 x – 0.04 | y = 0.90 x – 0.05 | t = -0.84, p = 0.448 |
| | AD-rel | y = 1.12 x + 0.01 | y = 0.95 x + 0.02 | |
| novelty-SAME | HC | y = 1.01 x + 0.05 | y = 1.24 x + 0.36 | m: yFADE < AiA old; |
| | SCD | y = 0.98 x + 0.03 | y = 1.17 x + 0.36 | t = -5.16, p = 0.007 |
| | MCI | y = 0.90 x – 0.03 | y = 1.15 x + 0.35 | n: yFADE < AiA old; |
| | AD | y = 1.07 x + 0.11 | y = 1.12 x + 0.30 | t = -9.54, p < 0.001 |
| | AD-rel | y = 1.01 x + 0.04 | y = 1.22 x + 0.37 | |
| memory-FADE | HC | y = 1.06 x – 0.01 | y = 1.24 x – 0.06 | m: yFADE < AiA old; |
| | SCD | y = 1.06 x + 0.00 | y = 1.25 x – 0.05 | t = -6.69, p = 0.003 |
| | MCI | y = 1.06 x – 0.01 | y = 1.23 x – 0.04 | n: yFADE > AiA old; |
| | AD | y = 1.07 x – 0.01 | y = 1.14 x – 0.03 | t = 4.49, p = 0.011 |
| | AD-rel | y = 1.08 x – 0.00 | y = 1.23 x – 0.08 | |
| memory-SAME | HC | y = 0.99 x + 0.16 | y = 1.81 x + 1.62 | m: yFADE < AiA old; |
| | SCD | y = 1.00 x + 0.17 | y = 1.82 x + 1.61 | t = -27.13, p < 0.001 |
| | MCI | y = 0.89 x + 0.03 | y = 1.69 x + 1.41 | n: yFADE < AiA old; |
| | AD | y = 1.03 x + 0.18 | y = 1.70 x + 1.38 | t = -28.16, p < 0.001 |
| | AD-rel | y = 0.99 x + 0.17 | y = 1.82 x + 1.63 | |

**Table S5.** *Comparison of stability analyses using different reference samples.* Regression equations for comparisons of single-value fMRI scores using, as reference samples, young subjects from the original study against yFADE subjects (column “yFADE vs. young AiA”; see Figure S3) or older subjects from the original study (column “old AiA vs. young AiA”; see Figure S4). Coefficients of the regression line were statistically compared with paired t-tests across participant groups (column “Statistics”; 4 degrees of freedom). Abbreviations: FADE, SAME, HC, SCD, MCI, AD, AD-rel: see Figure 2; y = dependent variable (here: scores using yFADE reference or old AiA reference), x = independent variable (here: scores using young AiA reference), m = slope, n = intercept.

### Supplementary Figures

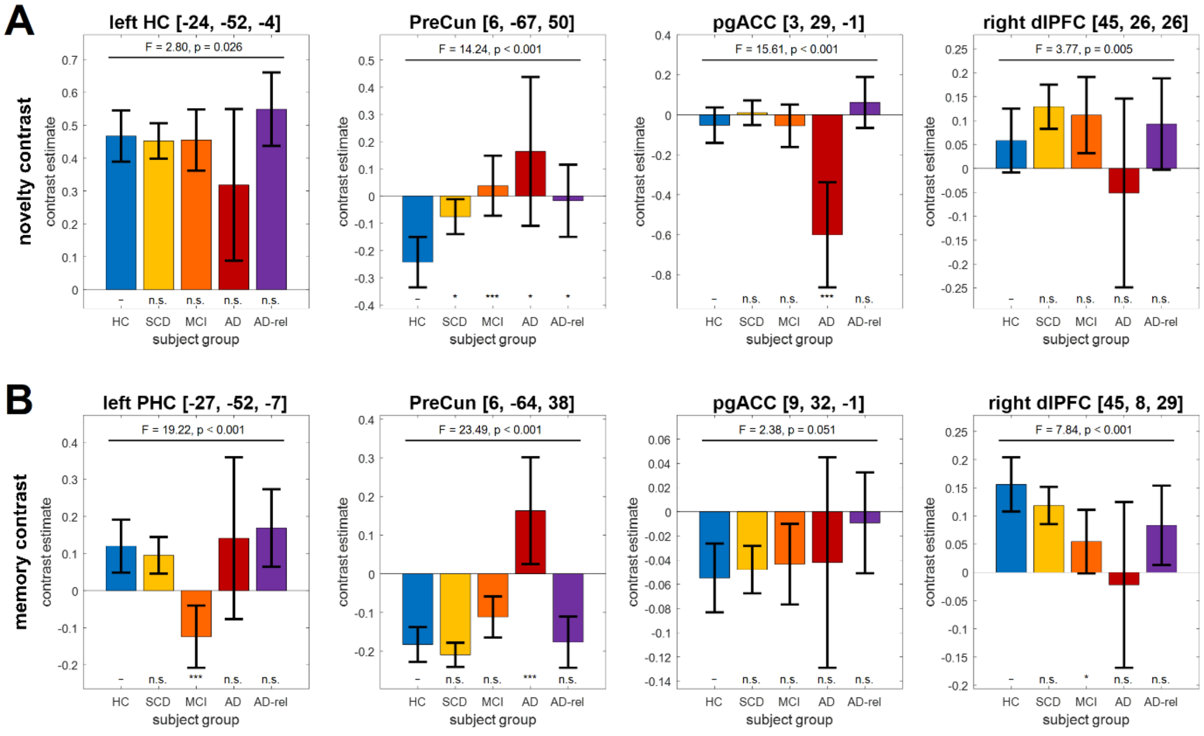


**Figure S1.** *Activation patterns by diagnostic group for novelty and memory (replication).* Bar plots group-level contrast estimates and 90% confidence intervals for **(A)** the novelty contrast (novel vs. master images) and **(B)** the memory contrast (subsequent memory regressor). Coordinates for parameter extraction were exactly identical to those found in the original study (cf. Soch et al., 2021, Fig. 2). Statistics inside the panels correspond to a one-way ANOVA across diagnostic groups (F/p-value; all F-values are $F_{4,463}$ statistics) and two-sample t-tests of each group against DELCODE healthy controls (significance markers). Abbreviations: HC = healthy controls, SCD = subjective cognitive decline, MCI = mild cognitive impairment, AD = Alzheimer's disease, AD-rel = AD relatives. Significance: * p < 0.05, Bonferroni-corrected for ** number of tests per region (4) or *** number of tests and number of regions (4 x 4). This figure corresponds to Figure 2 from the original publication.

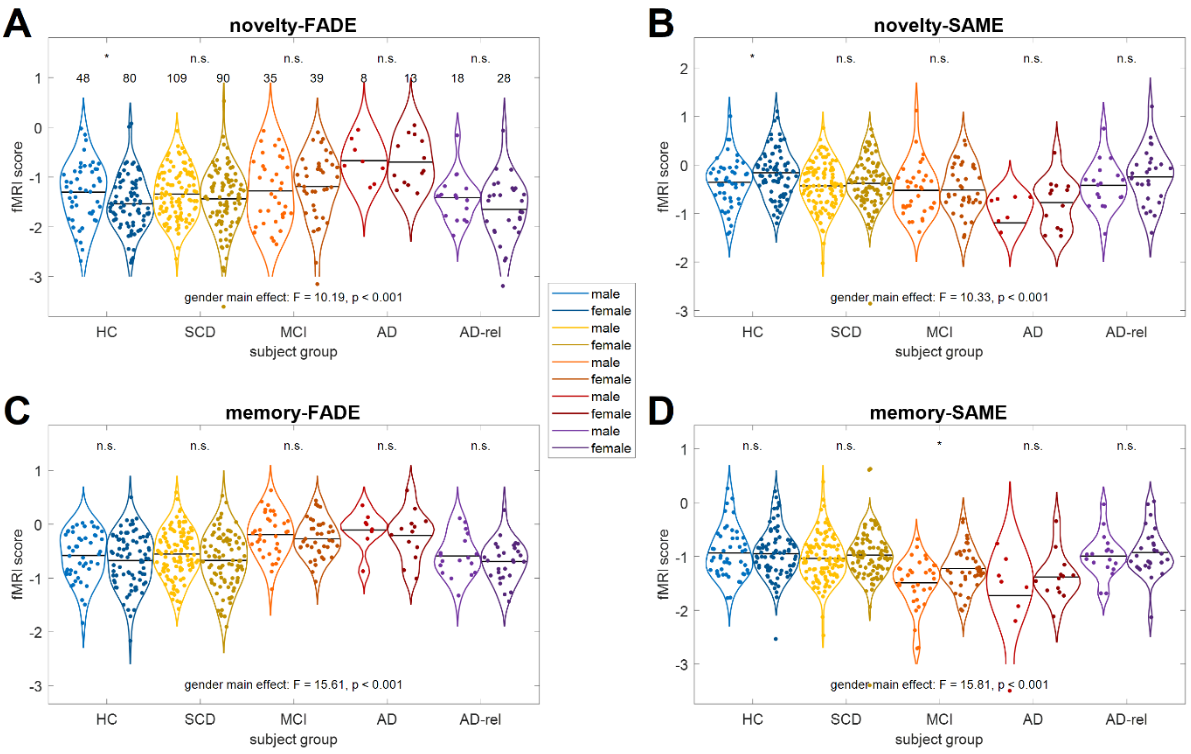


**Figure S2.** *FADE and SAME scores by diagnostic group and participant gender.* Single-value fMRI scores are shown for **(A)** the FADE score and **(B)** the SAME score computed from the novelty contrast as well as **(C)** the FADE score and **(D)** the SAME score computed from the memory contrast. The layout follows that of Figure 2. Sample sizes are given in the upper-left panel. Horizontal bars correspond to group-wise means. Markers on top of the x-axis denote a two-sample t-test between male and female participants (n.s. = not significant; * p < 0.05).

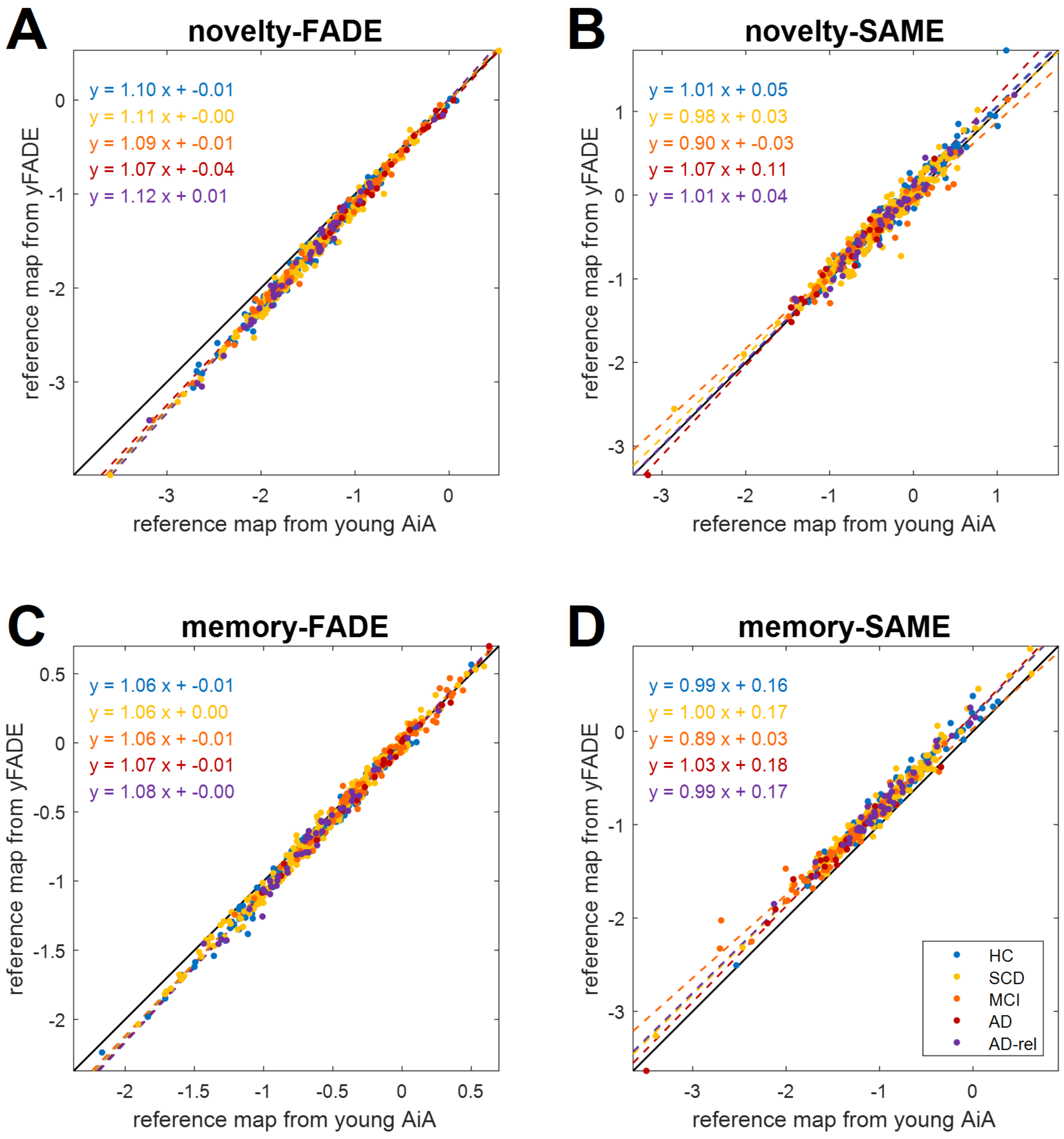


**Figure S3.** *Stability of the FADE and SAME scores as a function of reference sample (independent samples of young adults).* Comparison of fMRI scores of DELCODE participants, using reference maps obtained from either original reference sample of young subjects (young AiA) or the replication sample (yFADE), for **(A)** the FADE score and **(B)** the SAME score computed from the novelty contrast as well as **(C)** the FADE score and **(D)** the SAME score computed from the memory contrast. In all panels, the solid black line is the identity function, and the dashed black lines represent regression lines (equations given in top left). DELCODE participants include healthy controls (light blue), SCD patients (yellow), MCI patients (orange), AD patients (red) and AD relatives (violet). This figure corresponds to Figure 6 from the original publication.

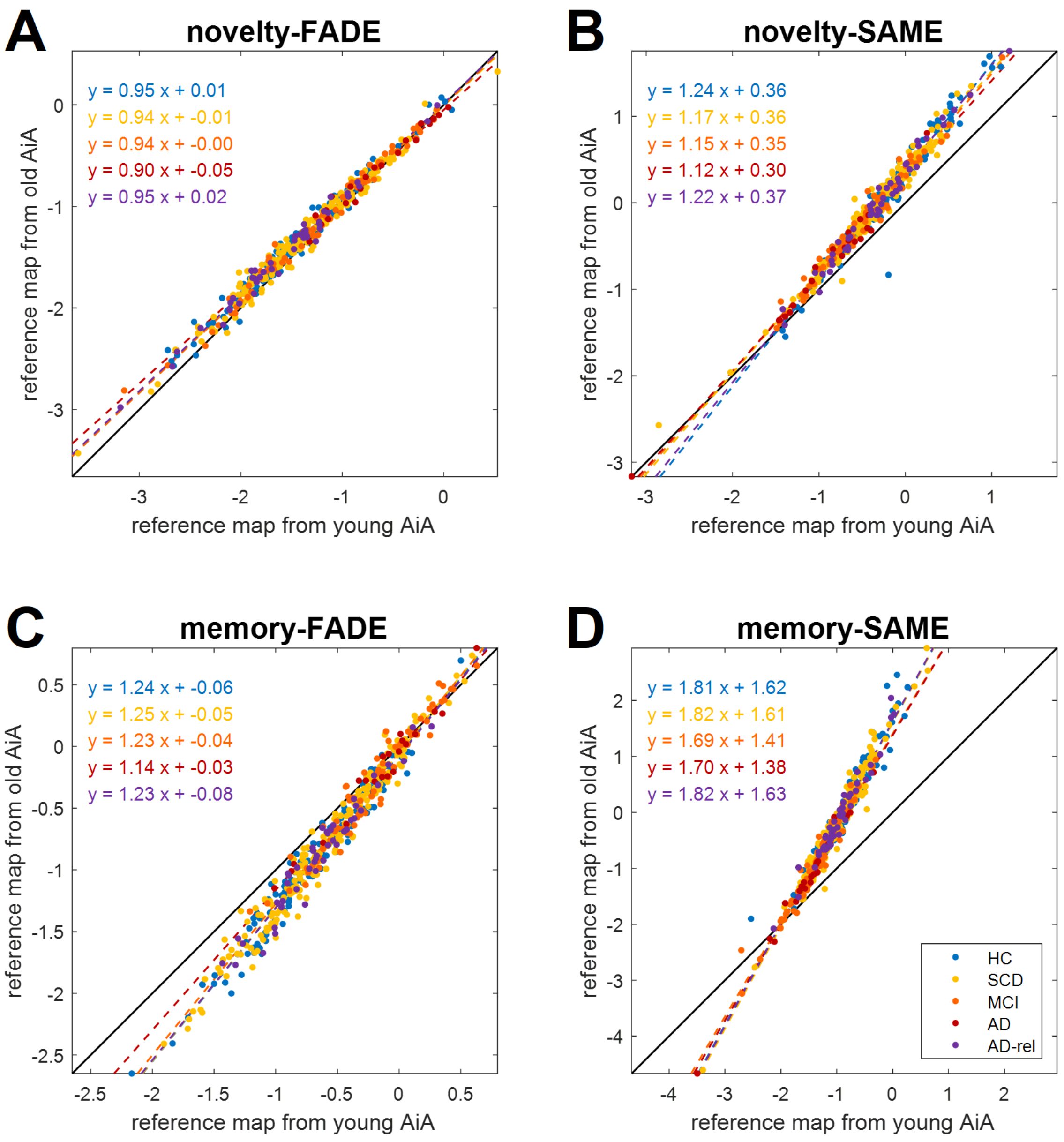


**Figure S4.** *Stability of the FADE and SAME scores as a function of reference sample (samples of young vs. older adults).* Comparison of fMRI scores of DELCODE participants, using reference maps obtained from either original reference sample of young subjects (young AiA) or older subjects (old AiA), for **(A)** the FADE score and **(B)** the SAME score computed from the novelty contrast as well as **(C)** the FADE score and **(D)** the SAME score computed from the memory contrast. In all panels, the solid black line is the identity function, and the dashed black lines represent regression lines (equations given in top left). DELCODE participants include healthy controls (light blue), SCD patients (yellow), MCI patients (orange), AD patients (red) and AD relatives (violet).

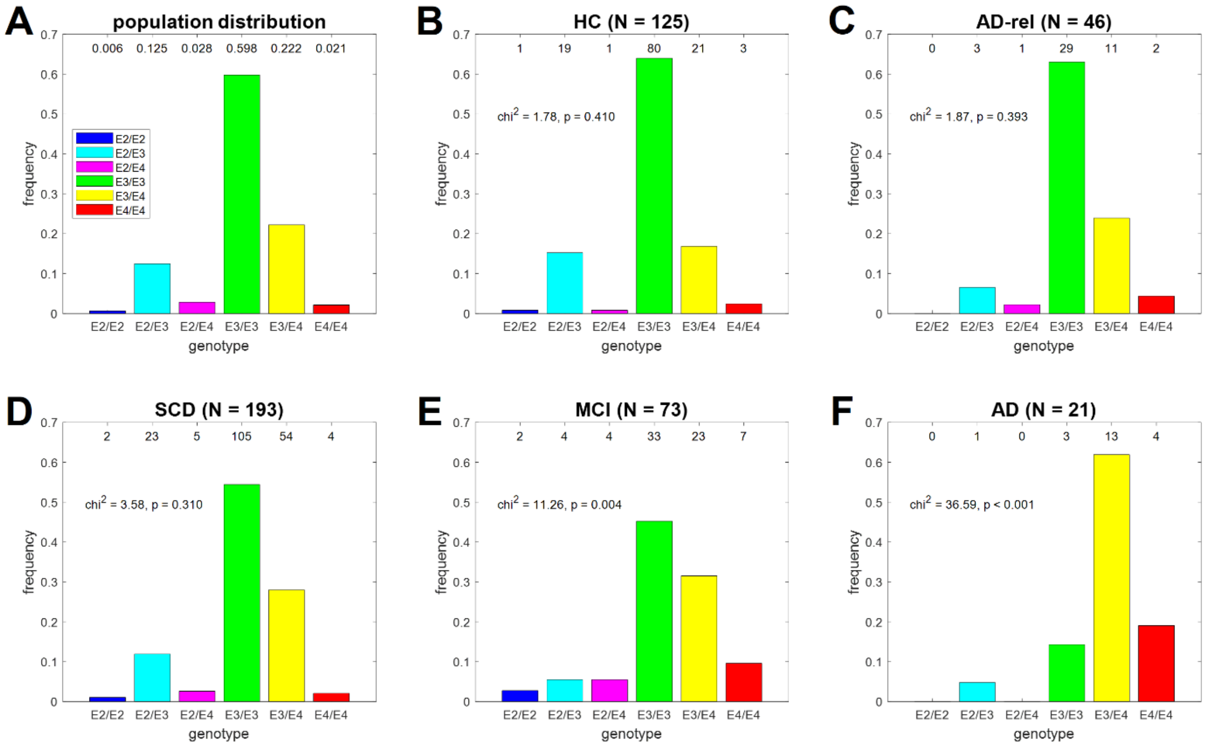


**Figure S5.** *Comparison of ApoE genotypes to population distribution.* Distribution of ApoE genotypes in **(A)** the general population, **(B)** healthy controls, **(C)** AD relatives, **(D)** SCD patients, **(E)** MCI patients and **(F)** AD patients. Frequencies of the population distribution were obtained from a behavioral genetics study in a comparable German sample (Li et al., 2019). When compared against the population distribution using a chi-squared goodness-of-fit test (2 degrees of freedom), a significant deviation was observed in MCI and AD patients. This figure corresponds to Figure S3A from the original publication.

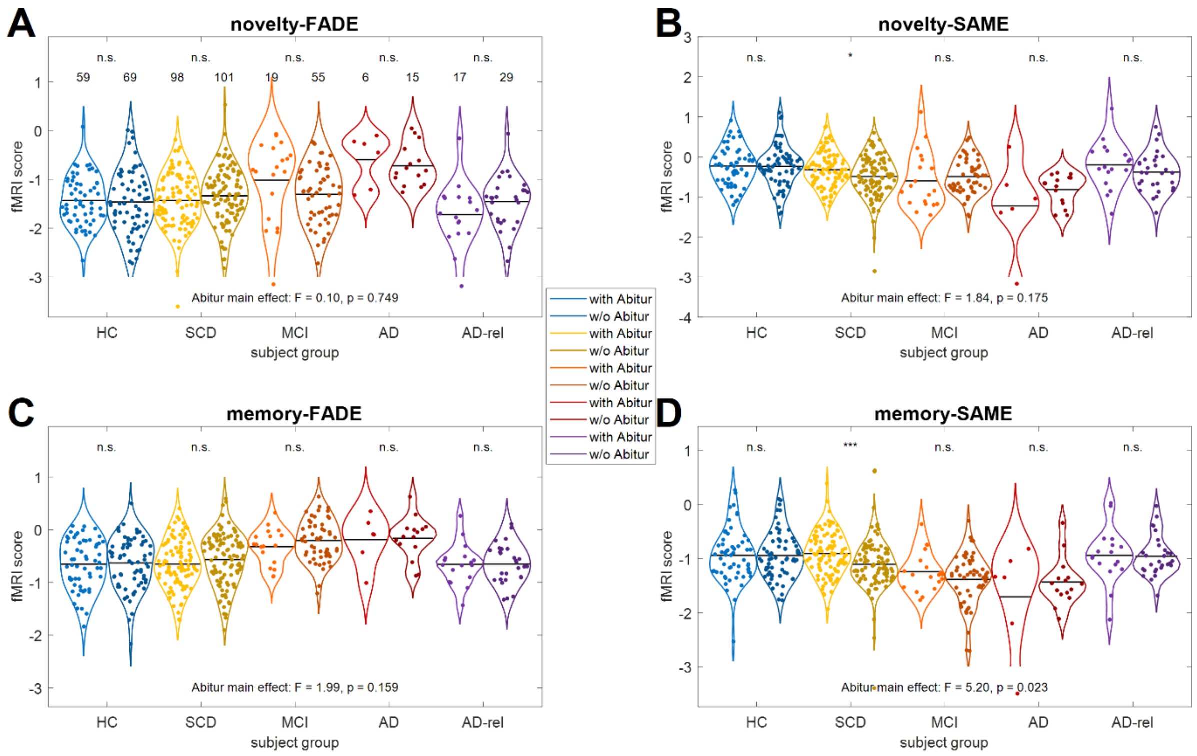


**Figure S6.** *FADE and SAME scores by diagnostic group and educational status.* Single-value fMRI scores are shown for **(A)** the FADE score and **(B)** the SAME score computed from the novelty contrast as well as **(C)** the FADE score and **(D)** the SAME score computed from the memory contrast. The layout follows that of Figure 2. Sample sizes are given in the upper-left panel. Horizontal bars correspond to group-wise means. Markers on top of the x-axis denote a two-sample t-test between subjects with and without Abitur (n.s. = not significant; * $p < 0.05$). This figure corresponds to Figure S4B from the original publication.

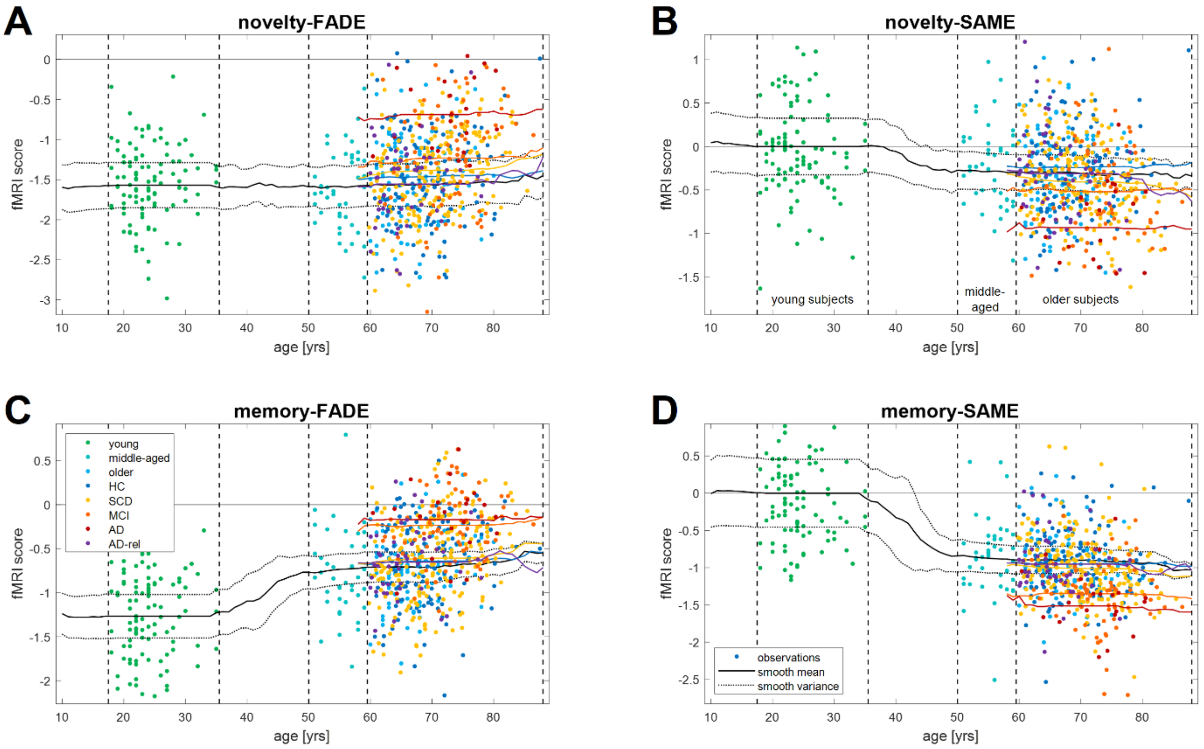


**Figure S7.** *FADE and SAME scores as a continuous function of age (including original study).* Single-value scores of all subjects were plotted against age (single dots) and smoothed using a sliding window of 32 years (solid lines) for **(A)** the FADE score and **(B)** the SAME score computed from the novelty contrast as well as **(C)** the FADE score and **(D)** the SAME score computed from the memory contrast. This display collects young (green), middle-aged (turquoise) and older (light blue) from the original study as well as healthy controls (dark blue), SCD patients (yellow), MCI patients (orange), AD patients (red) and AD relatives (violet) from the DELCODE study. The solid horizontal line in each panel represents zero, and the dotted lines correspond to the smoothed variance of subjects in the original study. This figure corresponds to Figure S5 from the original publication.

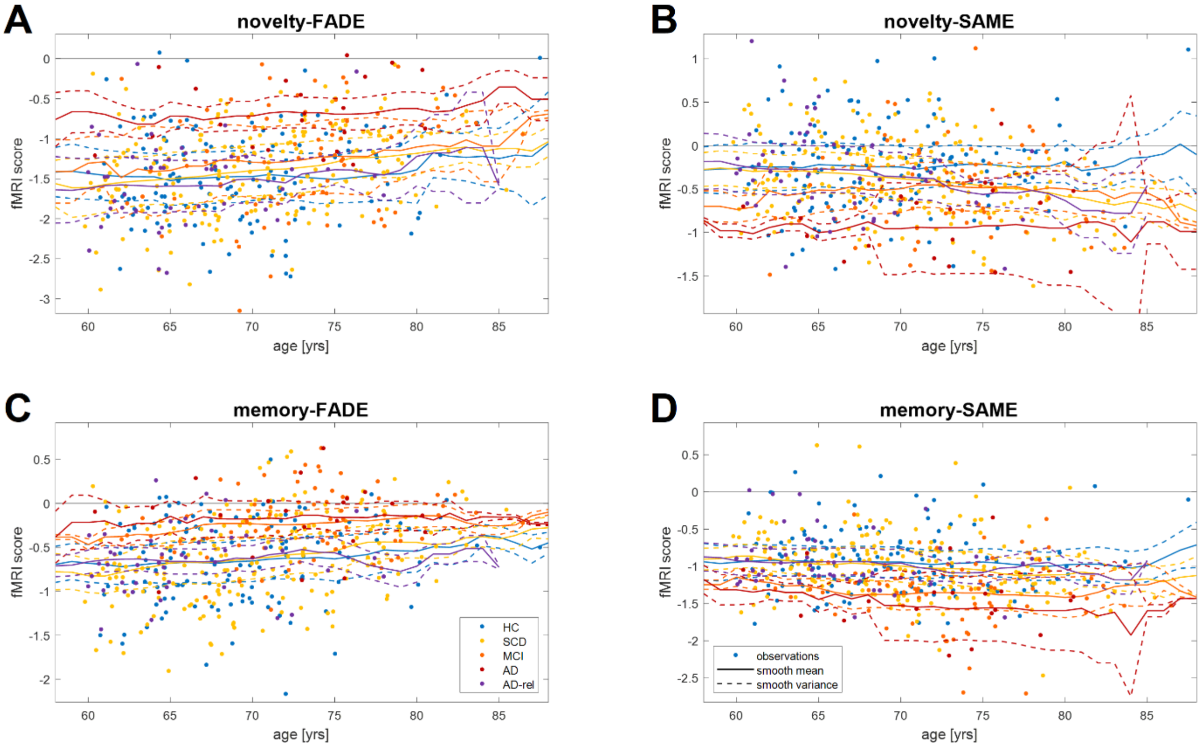


**Figure S8.** *FADE and SAME scores as a continuous function of age (excluding original study).* Single-value scores of all subjects were plotted against age (single dots) and smoothed using a sliding window of 16 years (solid lines) for **(A)** the FADE score and **(B)** the SAME score computed from the novelty contrast as well as **(C)** the FADE score and **(D)** the SAME score computed from the memory contrast. The solid horizontal line in each panel represents zero, and the dotted lines correspond to the smoothed variance of subjects in each participant group. This figure represents a zoom-in into the age range from ca. 60-90 years for the data shown in Figure S6.
